# An adaptive weight ensemble approach to forecast influenza activity in the context of irregular seasonality

**DOI:** 10.1101/2024.03.27.24304945

**Authors:** Tim K. Tsang, Qiurui Du, Benjamin J. Cowling, Cécile Viboud

## Abstract

Forecasting of influenza activity in tropical and subtropical regions such as Hong Kong is challenging due to irregular seasonality with high variability in the onset of influenza epidemics, and potential summer activity. To overcome this challenge, we develop a diverse set of statistical, machine learning and deep learning approaches to forecast influenza activity in Hong Kong 0-to 8- week ahead, leveraging a unique multi-year surveillance record spanning 34 winter and summer epidemics from 1998-2019. We develop a simple average ensemble (SAE), which is the average of individual forecasts from the top three individual models. We also consider an adaptive weight blending ensemble (AWBE) that allows for dynamic updates of each model contribution based on LASSO regression and uses decaying weights in historical data to capture rapid change in influenza activity. Overall, across all 9 weeks of horizon, all models outperform the baseline constant incidence model, reducing the root mean square error (RMSE) by 23%-29% and weighted interval score (WIS) by 25%-31%. The SAE ensemble only slightly better than individual models, reducing RMSE and WIS by 29%. The AWBE ensemble reduce RMSE by 45% and WIS by 46%, and outperform individual models for forecasts of epidemic trends (growing, flat, descending), and during both winter and summer seasons. Using the post-COVID surveillance data in 2023-2024 as another test period, the AWBE ensemble still reduces RMSE by 32% and WIS by 36%. Our framework contributes to the ensemble forecasting of infectious diseases with irregular seasonality.

**Significance statement:** In subtropical and tropical regions, irregular influenza seasonality makes accurate forecasting challenging. We test ensemble approaches using diverse statistical, machine learning, and deep learning models based on a unique multi-year surveillance record in Hong Kong. Performance of individual models varies by season and epidemic trend, but simple averaging ensemble cannot improve accuracy. Here we develop an adaptive weight ensemble approach, which updated individual model contributions dynamically. This approach halves the RMSE, outperforms all individual models in different settings and reducing RMSE by one-third even in post-COVID periods. Our method contributes to comparison and benchmarking of models in ensemble forecasts, enhancing the evidence base for synthesizing multiple models in disease forecasting in geographies with irregular influenza seasonality.

## INTRODUCTION

Influenza virus causes an estimated 3-5 million severe illnesses and 400000 deaths annually on average [1]. Forecasting infectious disease activity could inform public health response to outbreaks, such as preparing the increase of hospitalization [2, 3]. In regions with stable seasonality such as the continental U.S, forecasting of influenza and COVID-19 could be reliable, with establishment of forecasting hub [4] to generate ensemble forecast based on the forecast from several groups based on several mechanistic, statistical, machine learning and deep learning approaches [4–10].

In subtropical and tropical regions such as Hong Kong, influenza seasonality is irregular and causes difficulty. In these regions, the onset of influenza season is irregular, and there is potential of second influenza peak in summer seasons, so that accurate influenza forecast is challenging [11–13], particular the leverage of seasonality [4, 14]. Due to this challenge, modification of forecasting approach is required, and one previous study attempted to forecast the peak time and magnitude [13] but based on single model, while ensemble approaches have proven superior across a range of disease systems and geographies [4, 9, 15–18].

To overcome this challenge, we aim to develop a systematic approach to conduct nowcast, and 1-8 week ahead forecast of influenza activity in regions with irregular seasonality. Using Hong Kong as example, based on >20 years of data (1998-2019), we develop and evaluate several statistical, machine learning and deep learning approach. We further explore several ensemble forecast approaches that previously show superior performance in other diseases [4, 9, 17, 19] or geographics [20, 21]. We propose the idea of allowing model weights to be updated dynamically over time to better capture the rapid change of influenza activity in Hong Kong. Our approach framework and ensemble contribute to the evidence base of forecasting infectious diseases in geographies with irregular seasonality.

## RESULTS

### Influenza epidemics in Hong Kong

In Hong Kong, an indicator that combines influenza-like-illness intensity (ILI+) with the proportion of respiratory specimens positive for influenza each week is considered the gold standard to measure the intensity of influenza activity [22]. Over the twenty-one years from 1998 to 2019, there were 32 epidemics, in which 20 occurred in winter (November to April) and 12 occurred in summer (May to October). Epidemics have consistently occurred in winter throughout the study period, except for the year 2013 and 2016. In addition, the start week of epidemics is variable (Figure 1), with the started weeks of winter and summer epidemics ranging from week 5 (Nov, 29th)–21 (Mar, 21st) and week 27 (May, 7th)-47 (Sep, 18th) respectively.

**Figure 1:**
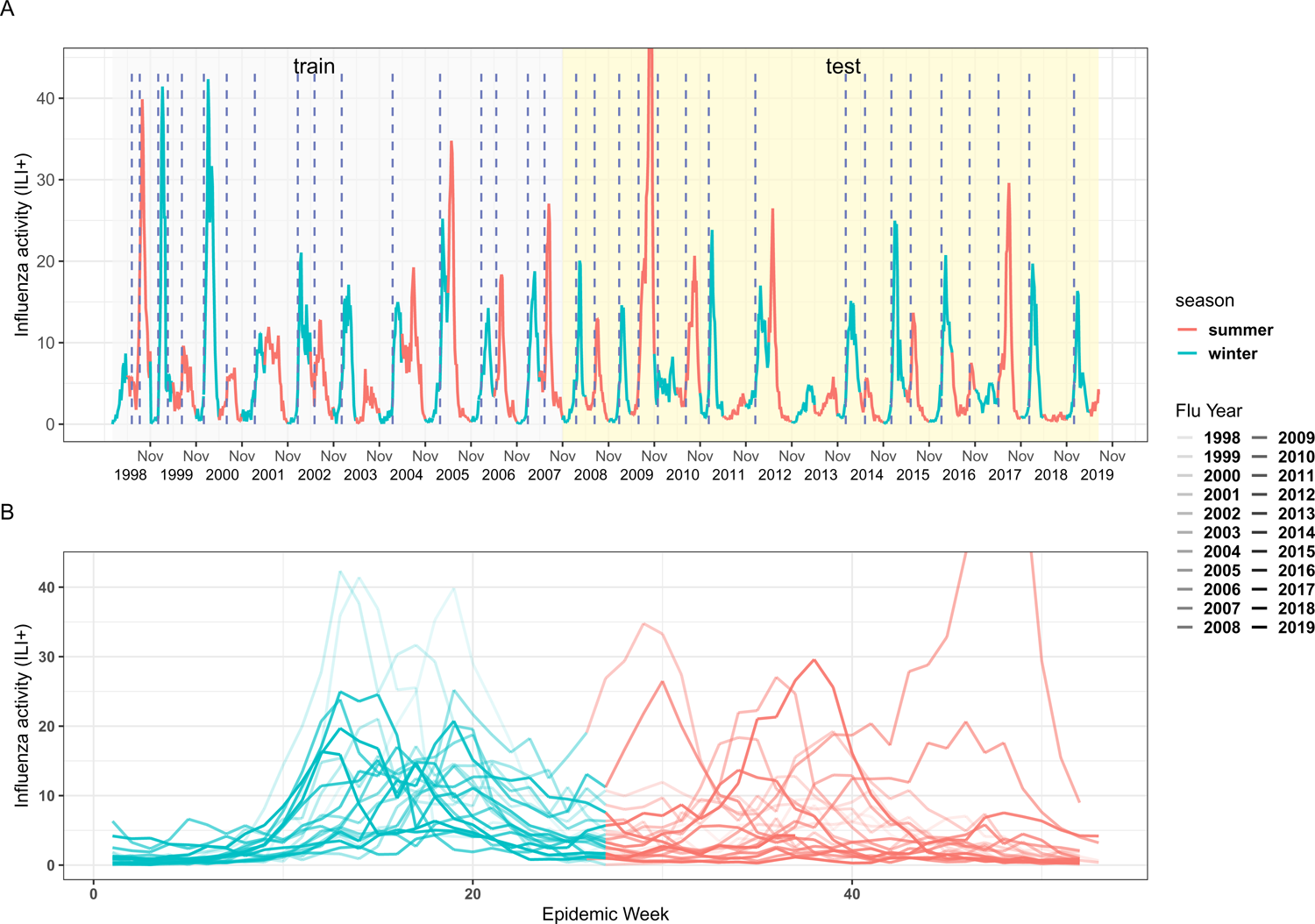
Influenza activity in Hong Kong. . Panel A: Influenza activity (ILI+) in Hong Kong from 1998 to 2019, encompassing the influenza season, and the training and testing periods. Blue dotted vertical lines indicate the start of epidemics. Panel B: Influenza trends in a year Hong Kong. Epidemic week is defined as November of the preceding year to the October in the current year.

There is a 1-week delay in the report of ILI+ (ie, at current week t, ILI+ is only available up to week t-1). Here, we mimic a real-time analysis that considers these delays to generate nowcasts and up to 8-week ahead forecasts. In 2009, the sentinel surveillance system was affected by the establishment of special Designated Flu clinics (not part of the sentinel network), in response to the pandemic influenza H1N1 outbreak. Therefore, the value of ILI+ is inflated [23], and we exclude 2009 forecasts in all evaluation.

### Overview of nowcasting and forecasting of ILI+

Given the irregular seasonality of influenza activity in Hong Kong, and the potential for influenza activity in summer, we generate nowcasts and forecasts of ILI+ throughout the year. This approach differs from season-based forecasting methods, which only forecast influenza activity from December to May in regions with regular influenza seasonality in winter and no influenza activity in summer.

We first develop a variety of statistical models to nowcast and forecast ILI+ up to 8 weeks ahead, incorporating epidemiological predictors including the past ILI+ values in the previous 14 weeks, the week and month of the year to reflect influenza seasonality, and meteorological data including weekly temperature, the range of temperature, absolute and relative humidity, rainfall, solar radiation, wind speed, and atmospheric pressure. Then the predictions from these individual models are combined into a single ensemble forecast. Eight models are tested, including the Autoregressive Integrated Moving Average model (ARIMA), Generalized Autoregressive Conditional Heteroskedasticity model (GARCH), Random Forests model (RF) [24], Extreme Gradient Boosting model (XGB) [25], Long Short-Term Memory Network Model (LSTM) [26], Gated Recurrent Units Network Model (GRU)[27], a Transformer-based time series framework named TSTPlus (TSTPlus) [28] and an ensemble of deep Convolutional Neural Network (CNN) models called InceptionTime Plus Model [29].

We use Jan 1998 - Oct 2007 as the training period, and Nov 2007-July 2019 as the test period. Regarding meteorological predictors, to avoid collinearity among them and determine the optimal lag, we compute the Pearson correlation between the ILI+ data and the predictors with varying lags during the training period to select 5 out of 8 meteorological predictors (see methods). Epidemiological predictors are always included in all models. Individual models are fitted on the selected predictors and observed ILI+ data, to generate forecasts in a rolling method: At each week t during the test period (Nov 2007-Jul 2019), we use the meteorological data up to week t and the ILI+ data only up to week t-1 to make forecast for the period t up to t+8. For statistical models (ARIMA and GRACH), the models are re-trained each week with most recent available data. For other models (machine learning and deep learning model) are only re-trained in the first week of November in each year with most recent available data, since weekly update required higher training cost with no improvement in performance (Figure S1).

Then, we generate ensemble forecasts based on individual models by using a simple averaging of the top 3 models and a model blending method that weights models based on past performance [18, 30, 31]. We also introduce a time adaptive decay weighting scheme, where performance with more recent data contributes more heavily to the weight of each model. For comparison with the ensemble and individual models, we consider a baseline “constant” null model, in which the ILI+ from weeks t to t+8 weeks remains the same as ILI+ at week t-1.

### Prediction intervals

Most machine learning and deep learning approaches do not provide prediction intervals, and our trials of using Monte Carlo Dropout (MCDropout) demostrate that MCDropout may generate worse point forecasts compared those without MCDropout (Figure S2). To address this limitation, we extend the previous approach using normal distribution with the point forecast as a mean, and the SD calculated by using the residuals from a rolling 20-week window [32]. Instead of fixing 20-week, we use the training data to determine the optimal week number for the rolling window for each prediction horizon and for each model (see method).

### Evaluation metrics

The performance of individual and ensemble models is evaluated and compared in the test period (Nov 2007 – Jul 2019). We primarily use the root mean square error (RMSE), symmetric mean absolute percentage error (SMAPE), and weighted interval score (WIS) to compare models. We also provide mean absolute error (MAE) and mean absolute percentage error (MAPE).

### Performance of individual models in nowcasting and forecasting

Eight individual models are used to forecast ILI+ up to a time horizon of 8 weeks. Most of these models could broadly capture the dynamics of ILI+ (Figure S3, Figure S4). Overall, across all 9 weeks of horizon (t=0 to t+8), all models could outperform the baseline constant model, based on RMSE (Figure 2), reducing the RMSE by 23%-29%, SMAPE by 17%-22% and WIS by 25%-31%. As the prediction horizon grew longer, the improvements of individual models become more apparent. For instance, all models outperform the baseline by reducing the RMSE by 22%-31% for 4 weeks ahead forecast and 33%-37% for 8 weeks ahead forecast (Table 1).

**Figure 2:**
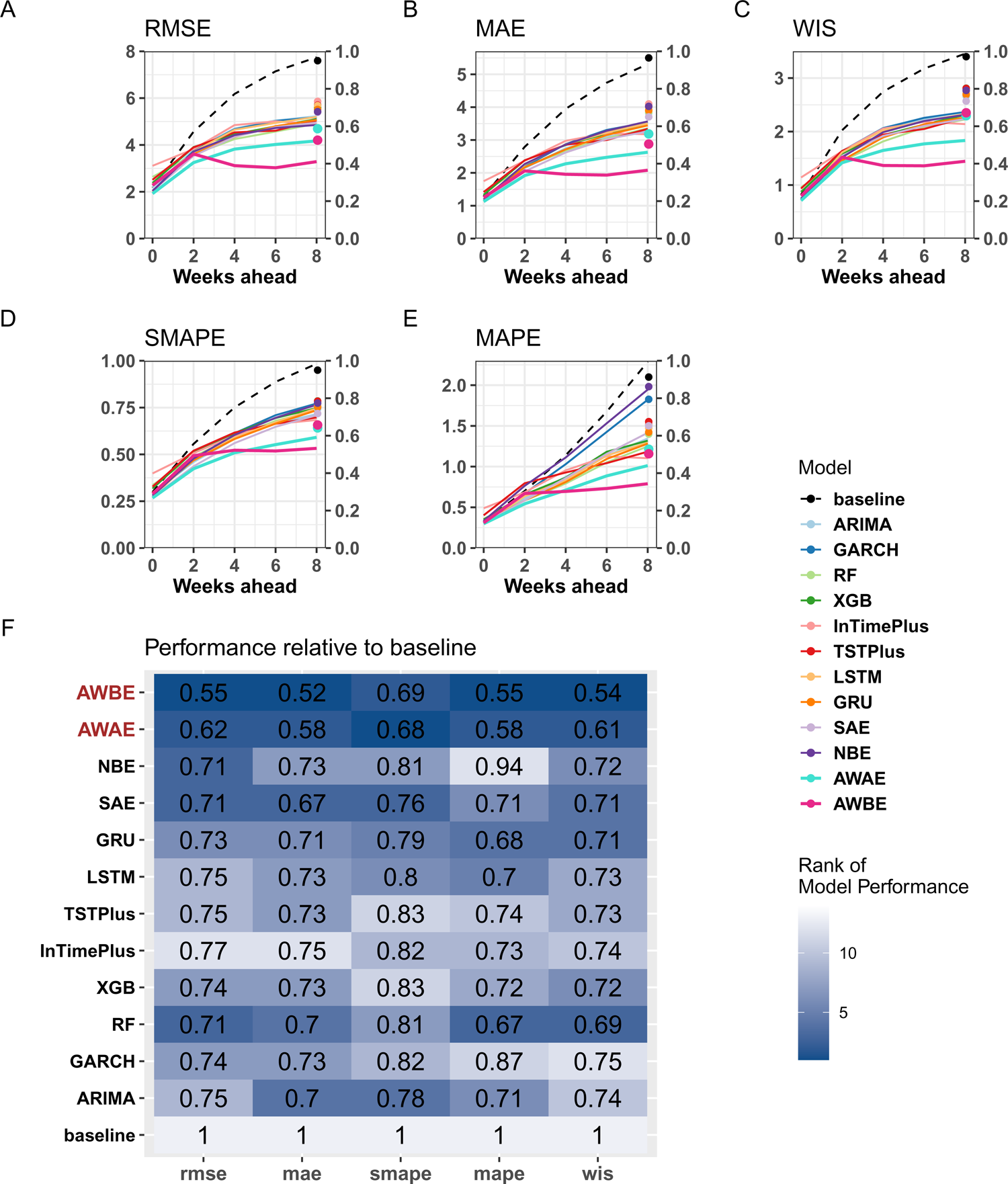
Performance comparison of individual and ensemble models over the testing period by prediction horizon. Panel A: RMSE. Panel B: MAE. Panel C: WIS. Panel D: SMAPE. Panel E: MAPE. Panel F: Relative performance of metrics compared to the Baseline. **Models**: ARIMA - Autoregressive Integrated Moving Average Model; GARCH - Generalized Auto-Regressive Conditional Heteroskedasticity Model; RF - Random Forest; XGB - Extreme Gradient Boosting; InTimePlus - InceptionTime Plus Model; LSTM - Long Short Term Memory Network; GRU - Gated Recurrent Neural Network; TSTPlus - Transformer-based Framework for Multivariate Time Series Representation Learning Model; **Metrics**: SAE - Sample Average Ensemble model; NBE - Normal Blending Ensemble model; AWAE - Adaptive Weighted Average Ensemble model; AWBE - Adaptive Weighted Blending Ensemble model.

**Figure 3:**
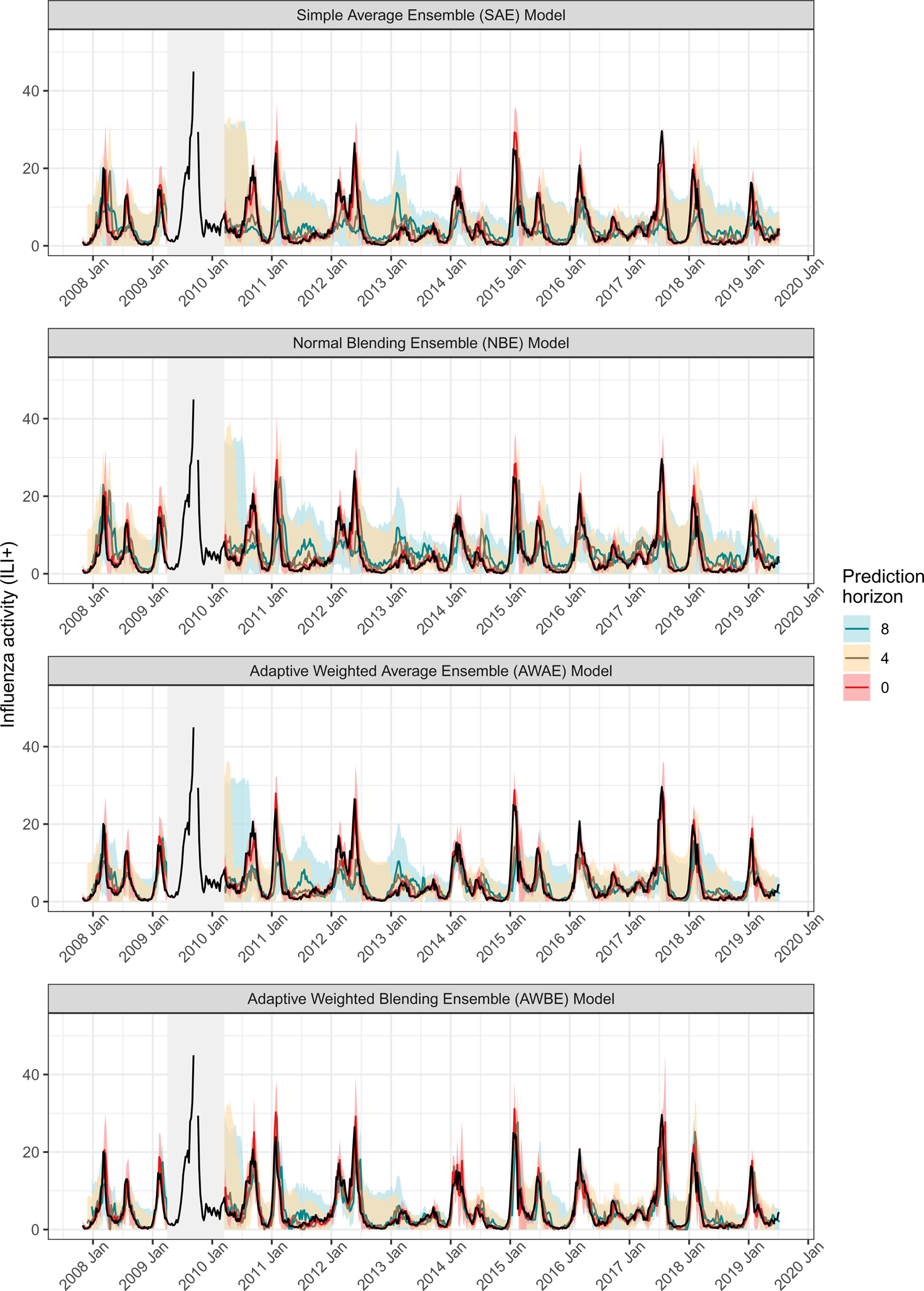
Trajectory of the forecasts of ensemble models. Red, yellow and blue indicate the point forecasts of 0 week, 4-week and 8-week ahead, accompanying the 90% prediction interval by shaded area of corresponding colors.

Next, we assess the performance of various models during distinct epidemic phases (Figure 4). The model with the best performance among phases is different, in which LSTM, RF and ARIMA performed the best in the growth, plateau and decline phase respectively, with reducing RMSE by 29%, 21% and 52% compared with the baseline model. We evaluate the model performance by season (winter or summer) (Figure 4), and find that the best-performing model in one season may not perform as well in the other. The RF model, which performs the best in winter, has the worst performance in summer among all individual models. Conversely, the InTimePlus model, which is the best in summer, has the worst performance in winter among all individual models. Since the relative skills of different types of models depend on the epidemic phase and the season, an ensemble would likely be well-positioned to improve overall performance.

**Figure 4:**
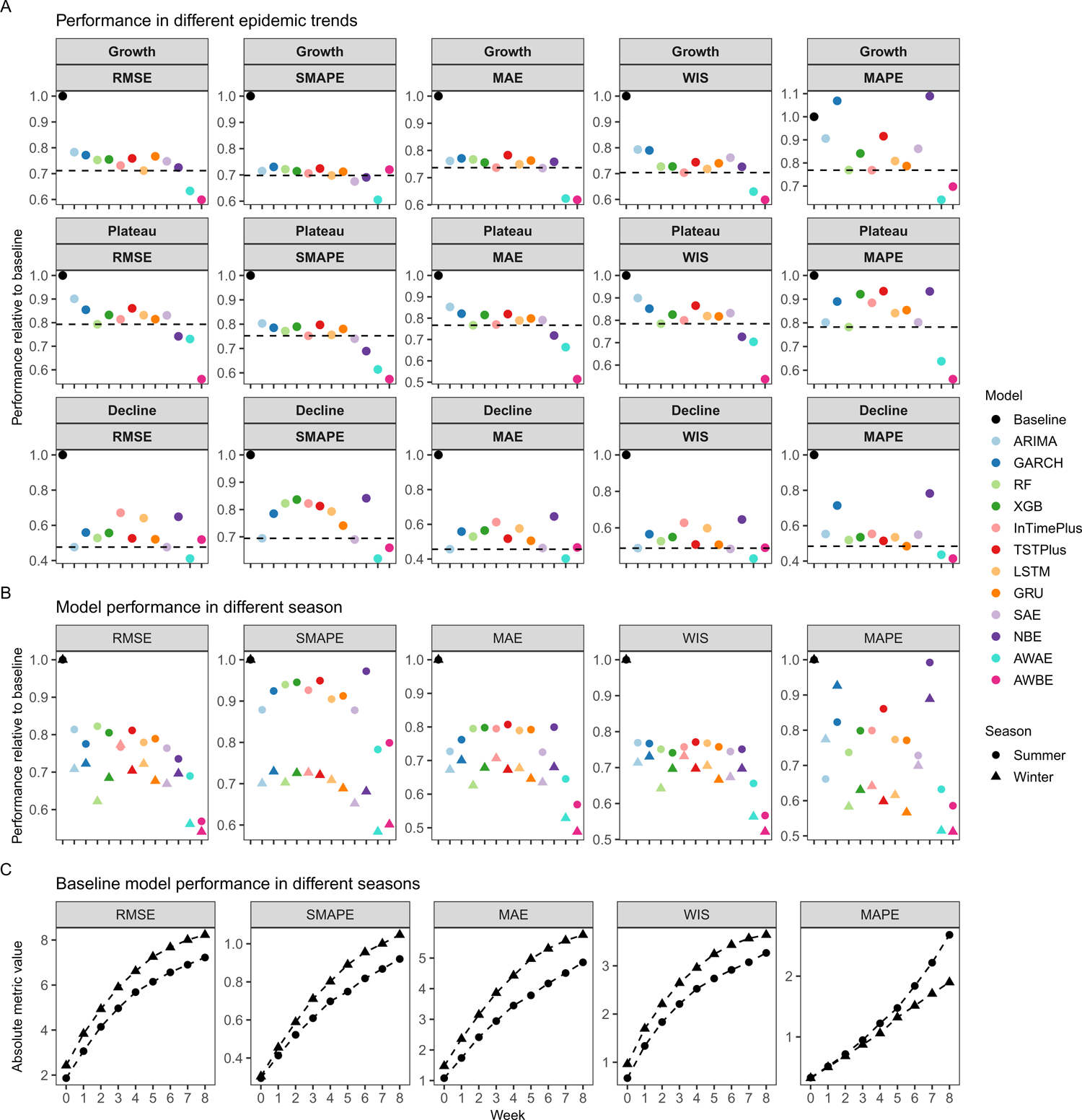
Model performance by epidemics phases and seasons. All metrics are relative to the Baseline model and include RMSE, SMAPE, MAE, WIS, and MAPE. Panel A: Model performance for distinct epidemic trends, with black dashed lines representing the best individual models. Panel B: Model performance by winter and summer seasons. Panel C: Baseline model performance by prediction horizons for summer and winter.

### Performance of ensemble models in nowcasting and forecasting

We first examine two ensemble approaches: the simple averaging ensemble (SAE) approach and the normal blending ensemble (NBE) approach. For the SAE model, we select the three best models at each week t based on RMSE calculated using data up to week t-1 and take the unweighted mean of these three individual forecasts. For NBE model, we fit a LASSO regression on observed ILI+ on predicted ILI+ from all individual models using data up to week t-1 and use the regression coefficients to assign weights in the nowcast and forecast periods. To further improve the forecast, we use an exponential time decay to weight the previous data, with the highest weight to the most recent week (t-1), when doing the abovementioned averaging or blending. These are named the Adaptive Weighted Average Ensemble (AWAE) model and the Adaptive Weighted Blending Ensemble (AWBE) model, respectively.

Compared to the baseline constant model, the SAE and NBE models reduced RMSE by 29%, SMAPE by 19-24%, and WIS by 28-30% respectively, which is slightly better than the best individual models. The performance further improved by considering decaying weights based on recent performance; the AWAE and AWBE ensemble models further reduce RMSE by 38% and 45%, reduce SMAPE by 31% and 32%, and reduce WIS by 40% and 46%, demonstrating that adaptive weighing could further improve the forecasting performance. Further evaluating the performance by different forecasting horizons, the improvement of AWAE and AWBE was more apparent at longer prediction horizons (Table 1, Figure 2&3). Furthermore, the AWAE and AWBE ensembles consistently improved forecast accuracy for different epidemic phases and for both summer and winter, compared to the SAE and NBE models or all individual models (Figure 4).

### Feature importance

The feature importance map illustrates the importance of different predictors for different models and time horizons, echoing model forecasting accuracy results and intuitive model interpretation (Figure 5). First, the ILI+ value in t-1 is highly important for all models, which is expected as this is the most updated information about the current epidemic state. Second, different models utilize the predictors in different ways. For statistical models (ARIMA and GRACH), except for ILI+ value in t-1, meteorological predictors and ILI+ value at t-2 and earlier are equally and weakly important, and there is little change in predictors across time horizons. For machine learning models (RF and XGB), predictors become more important when the prediction horizons are longer. Deep learning models have rather different feature importance map from the other approaches. For LSTM and GRU, the epidemiological predictors are more important than meteorological predictors. For InTimePlus and TSTPlus, only past ILI+ values are important in nowcast, but all other predictors become more important when the prediction horizons are longer. The differences in the learning capabilities on features and lags of different models also support the use of ensemble models and their improvement compared with individual models.

**Figure 5:**
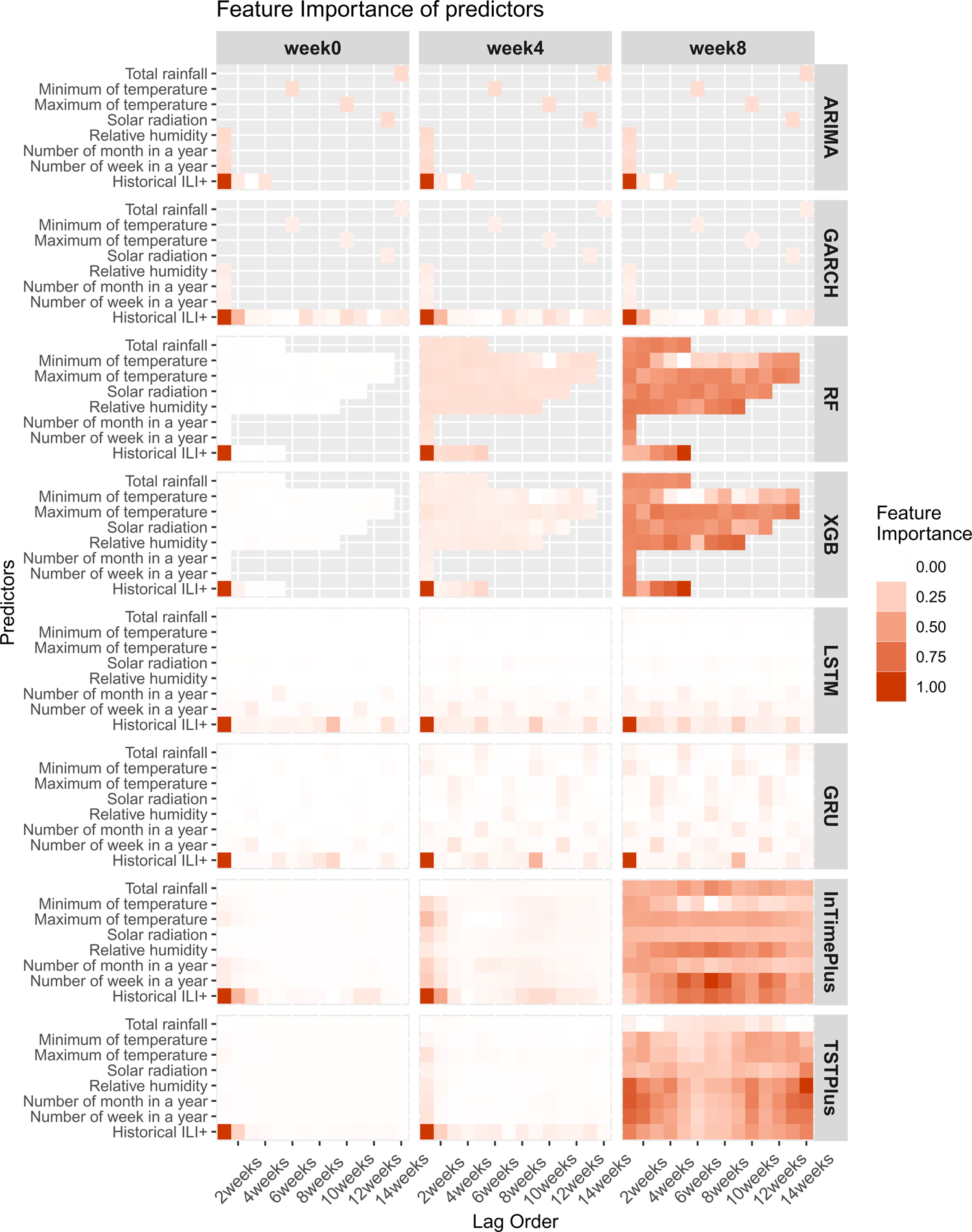
Feature importance in nowcast, 4-week, and 8-week ahead forecasts. Importance is measured by average regression coefficients in ARIMA and GARCH models, average feature importance in RF and XGB models, average saliency maps for LSTM and GRU models, and average permutation importance for TSTPlus and InTimePlus models.

### Requirement of the amount of training data and associated computational cost

To determine the amount of training data required, we vary the amount of training data by setting durations of training period from 1 to 9 years to retrain the models, generate forecasts, and compute the RMSE (Figure S6). There were decreasing trends in RMSE when the training period increases, particularly for the deep learning models. Overall, 7 years of data are required to have a comparable performance with the models using all available data (relative RMSE < 20%).

We also evaluate the computational cost of each model, measured by the running time of model training, for predicting a single week’s ILI+. The running time of ensemble models is the sum of the running time of all individual models plus the running time of the ensemble. We find that RF and XGB have a low training cost regardless of the amount of training data (<1 second), while the TSTPlus and InTimePlus have a high training cost, increased from around 6 seconds for 1 year of training data, to around 100-150 seconds for 9 years of training data. Overall, one run of training costs at most 150 seconds for the complicated deep learning model. For ensemble models, since the run of all individual models is required, it requires around 330 seconds for 1 year.

### Model Performance after COVID-19 Pandemic

In 2020-2023, Hong Kong implemented substantial public health and social measures to suppress COVID-19 outbreaks, and therefore influenza activity reached zero with no influenza epidemics in these three years [33, 34]. Given the potential impact on influenza transmission dynamics with the presence of SARS-CoV-2 viruses, we test our approaches in the post-COVID-19 era, using March 2023 to January 2024 as another testing period.

Overall, all models except for ARIMA and GRU still perform d better than the baseline model during 2023-2024, but slightly worse than pre-COVID era (Table 2, Figure S8). Only 6 out of 8 models could improve predictions, reducing the RMSE by 5-17% compared to the baseline model (Table S2). Ensemble models could further improve the predictions, with NBE, AWAE and AWBE reducing the RMSE by 24%, 21%, and 32%, respectively, compared to the baseline (Table S2, Figure 6), but not for SAE.

**Figure 6:**
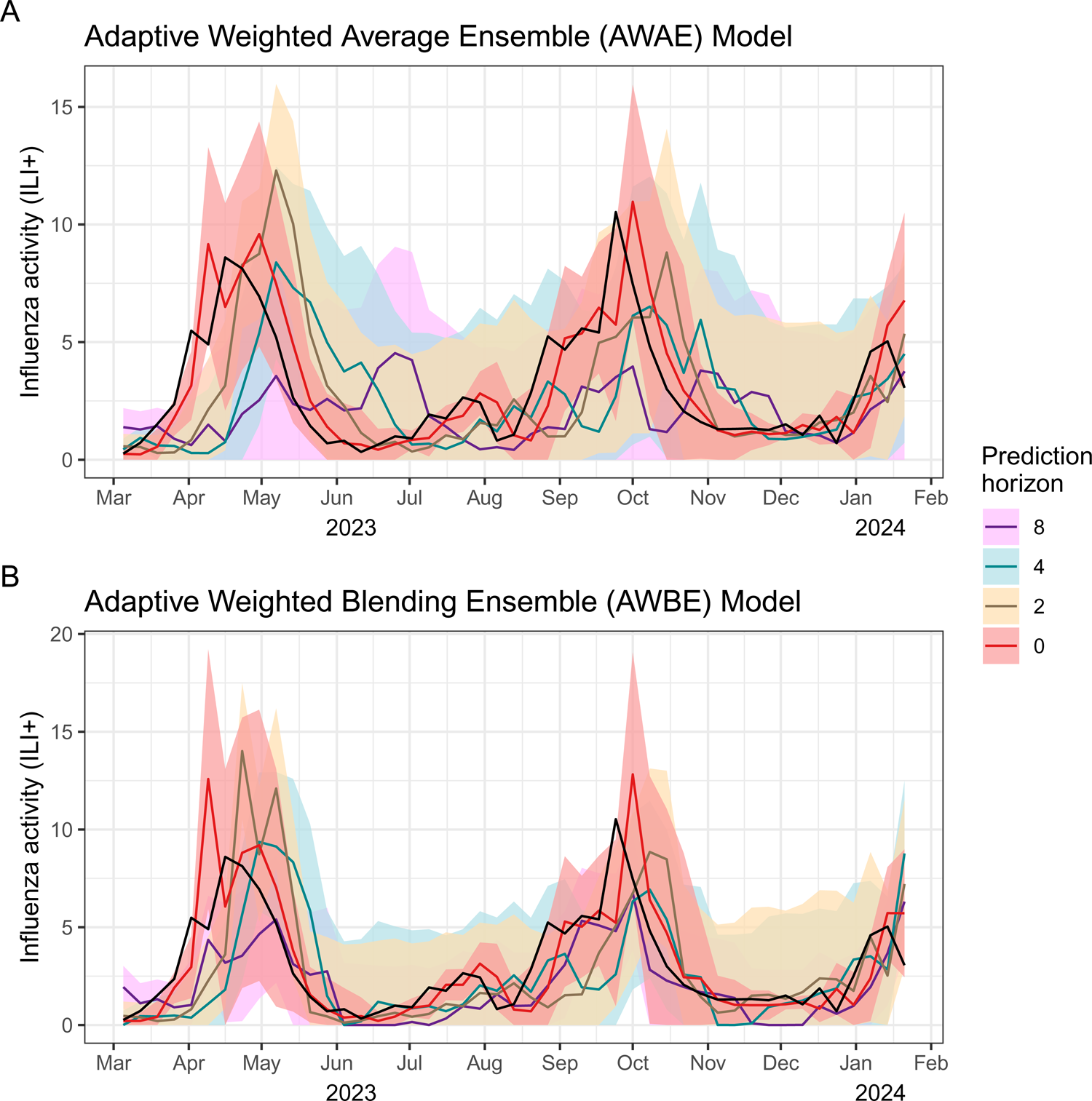
Trajectory of forecast of adaptive ensemble models during the post-COVID period. Red, yellow, blue and purple indicate the point forecasts of 0 week, 2-weel, 4-week and 8-week ahead, accompanying the 90% prediction interval by shaded area of corresponding colors.

## DISCUSSION

In this study, we evaluate the performance of various individual models and ensemble approaches to forecast the dynamic influenza epidemics (ILI+) in Hong Kong. Based on a unique dataset covering 32 epidemics in a subtropical region from 1998 to 2019, and using the last 11 years as the test period, we show that advanced machine learning and deep learning approaches can perform better than traditional time series models when making long-term forecasting. In evaluating ILI+ forecasts from current week to 8-week ahead, we find that the RF and InTimePlus models can reduce forecasting error (RMSE) by 27%, compared with the baseline “constant incidence” model. We further develop an adaptively weighted ensemble model that combines forecasts from individual models and reduces the RMSE by 45% compared with the baseline model. Overall, more sophisticated ensemble methods provide a greater edge over baseline and individual models at longer time horizons.

Influenza seasonality in Hong Kong is irregular, with varying outbreak onset times and potential for a second outbreak in summer, unlike in other regions such as the US and Europe. Consequently, we test various types of statistical, machine learning, and deep learning models. Compared to the baseline, the improvement in accuracy for these models in nowcasts is not readily apparent; however, it becomes more evident when the forecasting horizon is longer. We also find that different approaches are particularly well-skilled for different stages of an epidemic, or for summer/winter epidemics. For instance, RF performs best when forecasting ILI+ near the Plateau peak, while LSTM outperforms RF in the growth phase of epidemics. We investigate the feature importance (Figure 5) for individual models and the differences in learning capabilities concerning features. Most machine learning and deep learning approaches allow different types and orders of lagged variables to contribute to some extent, particularly for long-term forecasting, demonstrating the capability to capture the complex relationships between predictor factors and ILI+. These suggest the variations in the structures and learning capabilities of models may lead to performance differences in different seasons or phases of epidemics, motivating the need for ensemble approaches that can integrate skills from various models.

We test the simple averaging ensemble (SAE) that assigning same model weight to the top three models, and normal model blending ensemble (NBE) that allows for different model weights of all models, but they only slightly improve the accuracy compared with the best individual model by <1% in RMSE, even if the top three models in SAE and the model weights in NBE are updated weekly with same weight on all historical data in the estimation of model weights. This may suggest that using all historical data with equal weight in the estimation of model weights in ensemble may reduce the sensitivity to capture the change of ILI+ dynamics. This motivates the use of adaptive weight ensemble, which assigns higher weights to the most recent data points in estimating model weights to better capture the dynamics. Using this approach, the accuracy in the adaptive weighted averaging ensemble (AWAE) and adaptive weighted blending ensemble (AWBE) could improve the accuracy substantially, reducing 38% and 45% RMSE compared to the baseline, which is much higher than the 19% reduction in RMSE for SAE and NME. Our ensemble models AWAE and AWBE could still reduce the RMSE by 21% and 32% compared to baseline in March 2023 to February 2024, after 3-year of COVID-19 control measures in Hong Kong so that influenza activity was almost zero from 2020 to 2022. The slighted reduced performance may be due to the loss of population immunity against influenza for 3 years [34, 35] and the potential virus interference between influenza and SARS-CoV-2 [36]. Having more data in post-COVID era would be likely to improve the forecasting performance.

Unlike the well-developed influenza forecasting activity in regions with stable seasonality in winter like the US, there is limited forecasting work in subtropical regions like Hong Kong. To our knowledge, ours is the first study that attempted to forecast influenza activity at 0-8 week time horizons. Compared to a previous study that used a mechanistic model in Hong Kong [13], our ensemble forecast (AWBE) has a comparable performance in predicting the timing of the peak but a much higher accuracy in predicting peak magnitude (Figure S7).

Our systematic evaluation of different forecasting approaches has implications for advancing the science of infectious disease forecast. Machine learning and deep learning do not inherently provide probabilistic forecast. We test commonly used approach ‘Monte Carlo dropout’ to generate probabilistic forecast, but it worsens the accuracy of point forecast, in line with previous studies [37, 38]. Motivated by a previous approach [32], we use the training data to determine the optimal week number for the rolling window to compute the SD and generate the probabilistic forecast, and reduced the WIS by 40% and 46% for AWAE and AWBE ensemble model. Regarding the number of models in the ensemble forecast, we found that the optimal numbers could be different according to the evaluation metrics, so that in each forecasting task the optimal numbers of models in ensemble could be different, despite a previous analysis suggesting that four is the most optimal value in United States [39]. We compare frequency of retraining models and find that the accuracy from weekly and yearly retraining is similar. Despite retraining models weekly with most recent available information should have better performance intuitively, this may not be apparent in our study. With the higher computational cost for weekly retraining, our analysis suggests that yearly retraining could be a reasonable strategy.

Our study has some limitations. Our study is conducted retrospectively, which may tend to overestimate the performance of the forecasting models, compared to real-time forecasting. We do not consider other issues such as the delay of data release due to holidays, or data revision or correction, although they do not happen frequently in Hong Kong. Beside these limitations, we mimic real-time analysis by strictly utilizing data that would have been available at the time of forecast, to avoid hindsight bias. Second, we use regression coefficient, feature importance or the saliency maps to determine the importance of predictors for different types of models. Therefore, results from different comparison methods may not be directly comparable. Finally, we assume that influenza surveillance between 1998 and 2019 in Hong Kong is stable without important changes. While there are no documented changes except for 2009 pandemic outbreak, undocumented change may impact forecast accuracy.

In conclusion, we conduct systematic evaluation and analysis of various individual and ensemble models, and develop an adaptive ensemble approach that can halve the RMSE compared to a baseline constant model, in 0-8 week ahead forecast in Hong Kong, a region with irregular influenza seasonality. Our results demonstrate this approach can halve the forecast error compared to a baseline model, in 0-8 week ahead forecast. Our approach is feasible for comparisons and benchmarking of different prediction models, and integration of newly developed models in ensemble forecast, so that they can be applied to other infectious diseases in geographies with irregular seasonality.

## METHODS

### Data on influenza virus activity in community

In Hong-Kong, influenza activity in the general community is monitored through a sentinel surveillance network of outpatient clinics, which report the number of patients with influenza-like illness (ILI) each week. ILI is defined based on the presence of fever >37.8°C plus cough or sore throat. The rates of outpatient consultations for ILI per 10000 consultations are reported weekly. The public health laboratory also collects data on the weekly proportion of specimens from sentinel outpatient clinics and local hospitals that test positive for influenza virus. We generated a proxy measure of influenza activity (ILI+), derived as the weekly rates of outpatients with influenza-like illness multiplied by the weekly proportion of laboratory specimens positive for influenza virus, regardless of the subtype. We previously reported that this particular proxy provided a good indication of incidence of virus infection in the community based on hospital admissions [41].

We focus on the period from 1998 to 2019, before the outbreak of COVID-19. In 2009, the sentinel surveillance system was affected by the establishment of special Designated Flu Clinics (not part of the sentinel network) [23] (Figure 1). Therefore, the ILI+ value in that period is inflated and we exclude the data from Apr, 2009 to Mar, 2010 in our analyses.

### Overview of the modelling approach

We develop a framework to forecast the weekly ILI+ up to 8 weeks ahead. There is a one-week delay in availability of ILI+ data in real time so that in current week t, the most recent data available is ILI+ at week t-1. As a result, we develop forecasts for all weeks from t to t+8. We first develop a set of different statistical models to forecast ILI+. These individual forecasts are then combined into a single ensemble forecast based on different aggregation approaches. We divide our study into a training period (January 1998 to October 2007) and a test period (November 2007 to July 2019). The models are fitted during the training period and the hyperparameters are tuned based on performance over the same period. Then we conduct individual forecasting and model ensemble on a rolling basis over the test period. We evaluate the performance of both the individual and ensemble models.

### Individual forecasting models

In this article, we have established a total of 8 individual models, including the Autoregressive Integrated Moving Average model (ARIMA), Generalized Auto Regressive Conditional Heteroskedasticity model (GARCH), Random Forests (RF) [24], Extreme Gradient Boosting model (XGB) [25], Long Short-Term Memory Network model (LSTM) [26], Gated Recurrent Units Network Model (GRU) [27], a Transformer-based time series framework model (TSTPlus) [28] and an ensemble of deep Convolutional Neural Network (CNN) models which is called InceptionTime Plus Model [29] (Full details in appendix). We added a persistence model as a null baseline, in which the values of 0- to 8-week ahead forecast of ILI+ in week t are the same as the ILI+ value of week t-1 [9, 32].

For each individual model, we include a set of predictors that may inform the trajectory of ILI+. We consider two types of predictors: epidemiological predictors, including the week number and month of the year, which may capture influenza seasonality, and prior ILI+ values from week t-1 to t-14; and 8 meteorological predictors, including the maximum and minimum values of weekly temperature, the range of weekly temperature, absolute and relative humidity, rainfall, solar radiation, wind speed and atmospheric pressure.

Before the model fitting, we conduct a covariate selection for 8 meteorological predictors using Pearson’s correlation between each predictor and ILI+, and select 5 meteorological predictors with the highest correlations, including the maximum and the minimum of temperature, the relative humidity, total rainfall, and solar radiation. Then at each week t, the best lag is automatically selected for each predictor and model based on the Pearson correlation analysis between ILI+ data and the predictors at varying lags. Considering the differences in learning capabilities among models, the ARIMA and GARCH models only have a single predictor x_t-k_ with the optimal lag k for each selected predictor, while the RF and XGB models have the predictors from lag 1 to the optimal lags (x_t-1_, x_t-2_,-, x_t-k_). For deep learning model, for each select predictor, lag 1 up to lag 14 is included. In addition, influenza seasonality in Hong Kong is well documented (with a winter peak and summer peak) [42], so that week and month of year are included by default in all models.

### Model evaluation

For each week t in the test period, several evaluation metrics are computed using observed data from t to t+8. Model performance is evaluated primarily using the root mean square error (RMSE) [43], which has been widely used in previous literature. Furthermore, we also employ the symmetric mean absolute percentage error (SMAPE), a variant of the mean absolute percentage error (MAPE) with added symmetries to eliminate skewness and make the model resistant to outliers, and which is more appropriate when the ILI+ values are close to zero [44]. We also provide the mean absolute error (MAE) and MAPE for reference. Regarding probabilistic forecast, we employ the weighted interval score (WIS) as the evaluation metric. The WIS is a proper score that combines a set of interval scores for probabilistic forecasts that provide quantiles of the predictive forecast distribution. It measures how close the entire distribution is to the observation [45].

To assess the performance of interval prediction, we initially employed Monte Carlo Dropout (MCDropout), the most commonly used interval prediction method in deep learning [37, 38]. However, we found that MCDropout may reduce the accuracy of point forecast (Figure S2). Therefore, we adopted an interval prediction method grounded in Gaussian distribution proposed in Aiken et al. [32], with the assumption that the prediction follows a normal distribution characterized by a mean of y∼ and a standard deviation of σ. The method for calculating σ is outlined as follows: For a fixed calculation window length *k* and one given week *t*, we first compute the prediction errors (β = ў - y) for the historical *k* weeks, denoted as β_t-k-1_, β_t-k_, β_t-1_, and then calculate the standard deviation σ based on this sequence. Instead of setting k=20 in Aiken et al., we use the training data to determine the optimal k. We select the optimal k for each model by first testing k to be 5 to 50 and then calculating the 90% coverage rate for each prediction horizon (0-8 week) and each k. Based on this, we obtain the optimal window length for each prediction horizon and for each model, by selecting the k that the corresponding 90% coverage rate is closest to 90%. We then apply these optimal window lengths to the testing period and obtain the final interval prediction results.

We also evaluate the model performance by winter and summer seasons, and by trend of the epidemics. We define the trends to be growth (increasing for three consecutive weeks), decline (decreasing for three consecutive weeks) and plateau (the weeks between adjacent growth and decline phases).

### Model ensemble

Ensemble models are built to combine forecasts from individual models into a single forecast, where each individual forecast can be assigned a weight. Given a set of *m* models, denote rri as the forecast of model *i* at week *t* and *w^t^_i_* the ensemble coefficient for model *i* at week *t*, then the ensemble forecast Ў_t_ is:

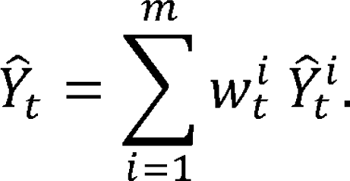

We test the following approaches to determine the weights: 1) Simple averaging ensemble (SAE) model: In this ensemble model, we select the top three models based on RMSE at each week t, and then take the unweighted mean of the individual forecast of these 3 models [9] (ie w^i^_t_ =0 if model *I* is not selected and 1 if it is). 2) Normal Blending Ensemble (NBE) model: In this approach, we fit a LASSO regression of the observed ILI+ up to week t-1 on the predicted ILI+ from different models to obtain the coefficients to generate ensemble forecast for week t [46].

We introduce adaptive weighting on the past ILI+ value, with higher weights for observations closer to the date of conducting forecast. We use the exponential time decay function to weight past ILI+ values to generate an ensemble forecast. For week t, the weight of the ILI+ of week t-k is:

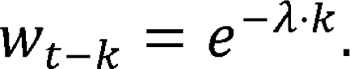

We set lambda to be 0.1, so that after around 1 year, the weight is <0.01. We use these weights in the simple averaging method when we compute the RMSE to select the best three models. We call this simple averaging method with time-decay the adaptive weighted average ensemble (AWAE) model. We also use a similar approach for the blending ensemble, in which we apply these weights when fitting the LASSO regression. This last approach is the adaptive weighted blending ensemble (AWBE) model.

### Implementation

The entire experiment was carried out on a Windows system furnished with a 12^th^-generation Intel i7 CPU processor and an Intel UHD Graphics 770 graphics card, to ensure fair comparison of model running time. Statistical analyses were conducted using Python version 3.9 and R version 4.0.5 (R Foundation for Statistical Computing, Vienna, Austria).

### Feature importance

For interpretability purposes and to better understand the role of each predictor, we analyze the feature importance of each individual model. Specifically, we obtain regression coefficients for each variable for regression-based model (ARIMA and GARCH) and feature importance for machine learning model (RF and XGB) [47]. For deep learning models, we use saliency maps for LSTM and GRU [48] and permutation importance for TSTPlus and InTimePlus [49, 50], based on their different model structures. As models are trained dynamically, the feature importance in each model may change from week to week. We normalize the importance data and take an average for the entire study period. Figure 5 compares the average feature importance of different predictors and models with different lag order for ILI+ nowcast, and 4 weeks and 8 weeks ahead forecast.

## Data Availability

All data produced in the present study are available upon reasonable request to the authors

## ACKNOWLEDGEMENTS

This project was supported by the Theme-based Research Scheme (Project No. T11-712/19-N) of the Research Grants Council of the Hong Kong SAR Government, and the Health and Medical Research Fund, Food and Health Bureau, Government of the Hong Kong Special Administrative Region (grant no. 21200292). BJC is supported by the AIR@innoHK program of the Innovation and Technology Commission of the Hong Kong SAR Government.

## AUTHOR CONTRIBUTIONS STATEMENT

T.K.T. designed research; T.K.T. and Q.D. performed research; Q.D. and T.K.T. contributed new analytic tools; T.K.T., Q.D. analyzed data; and T.K.T., Q.D., B.J.C. and C.V. wrote the paper.

---

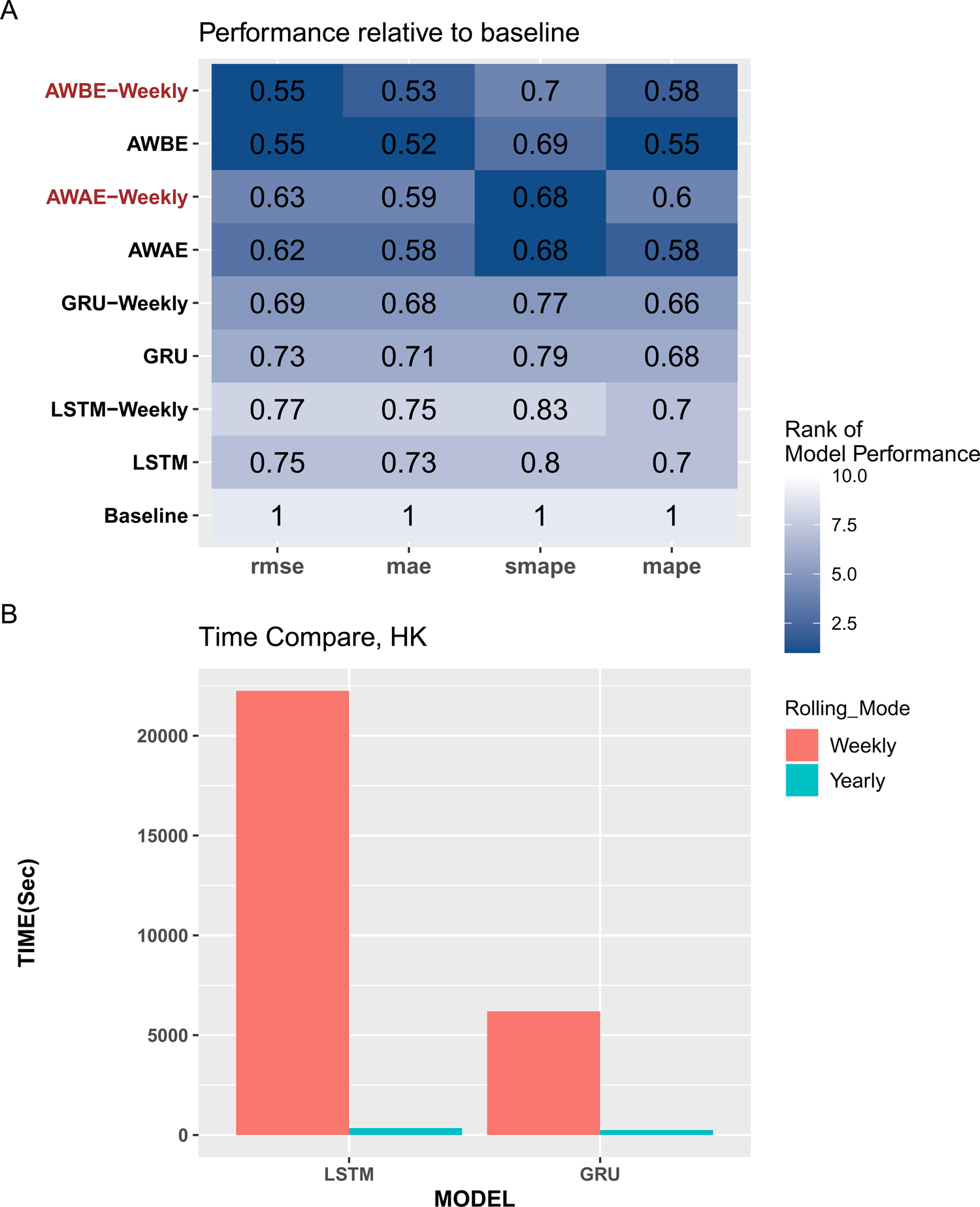

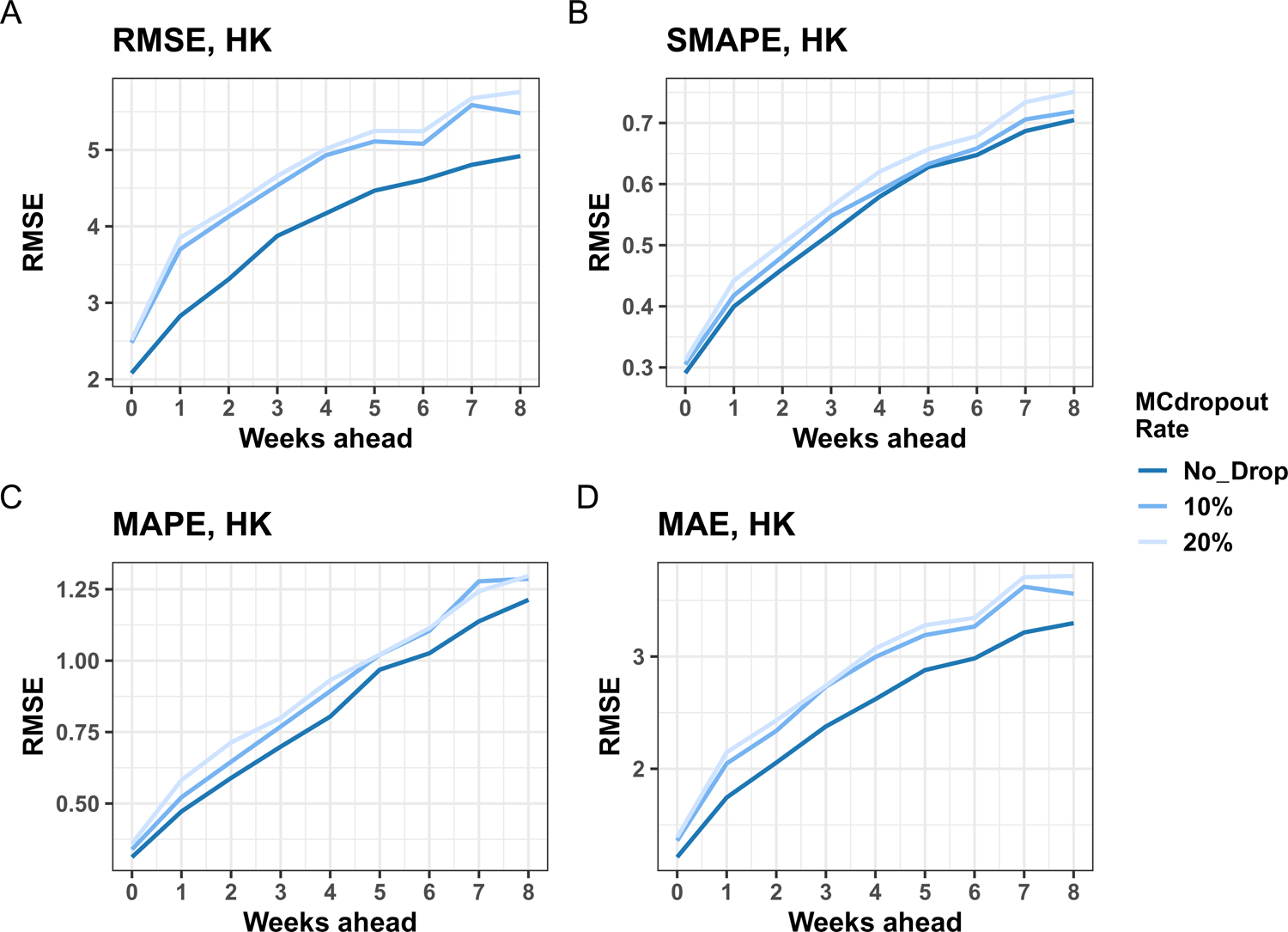

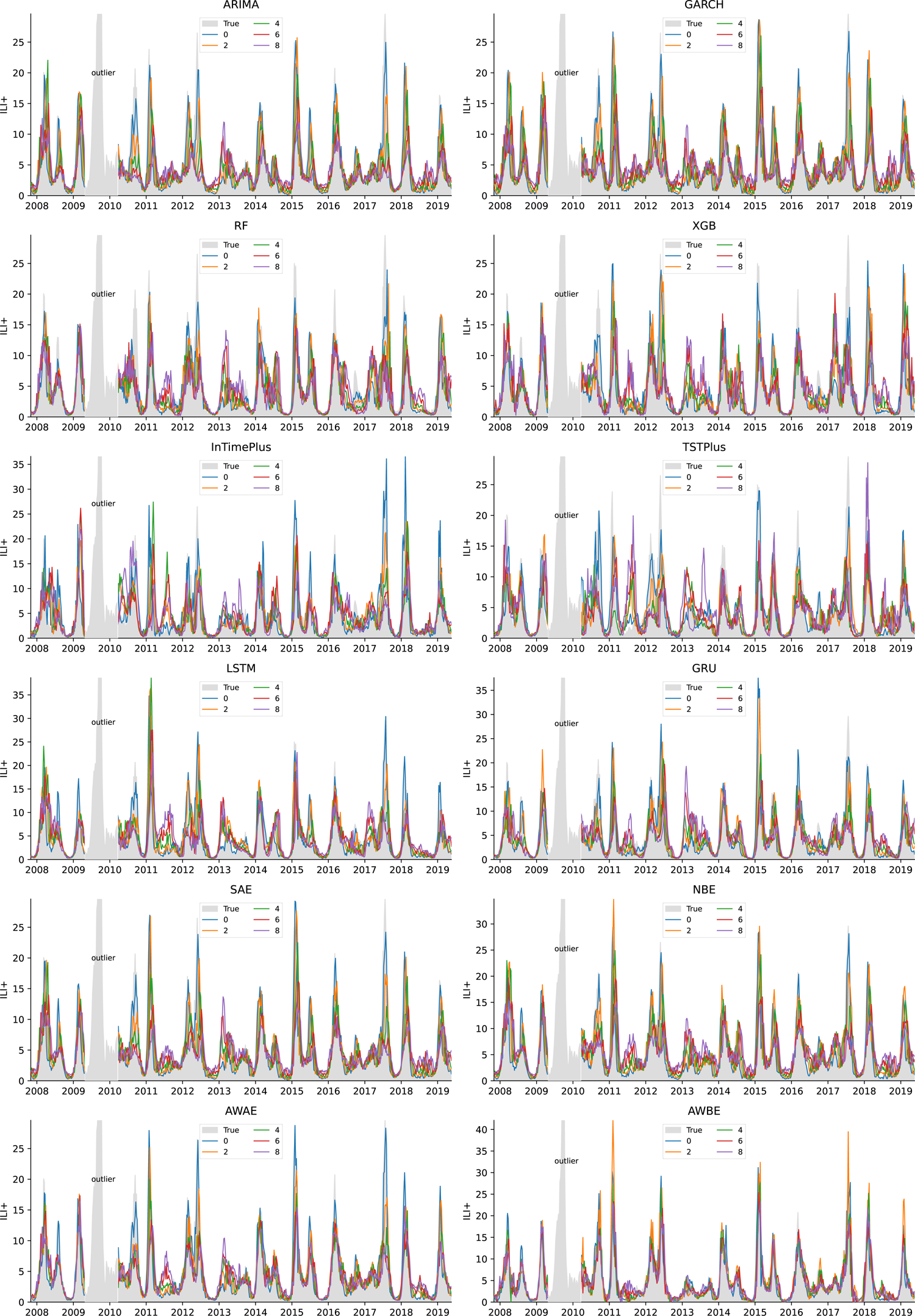

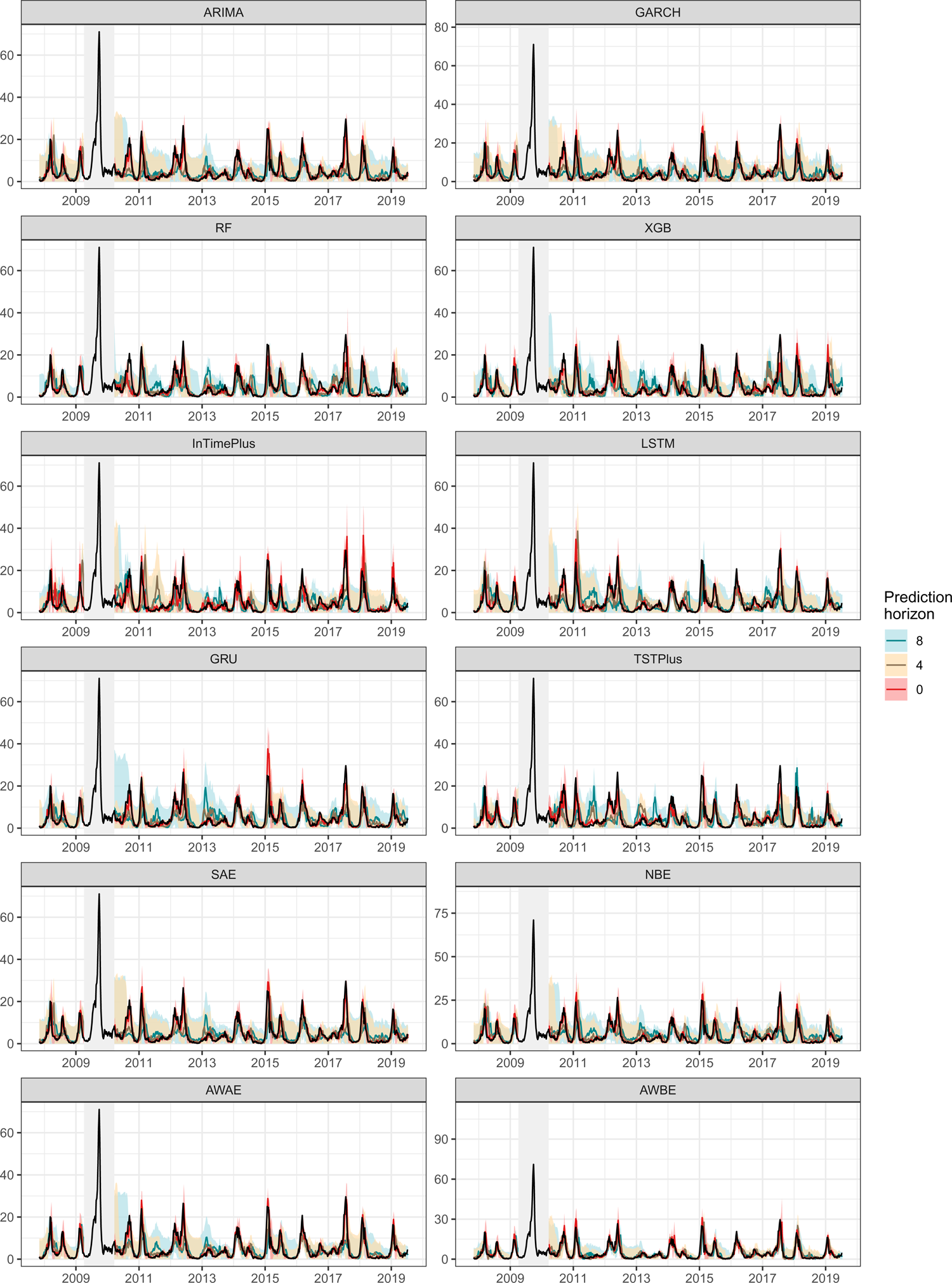

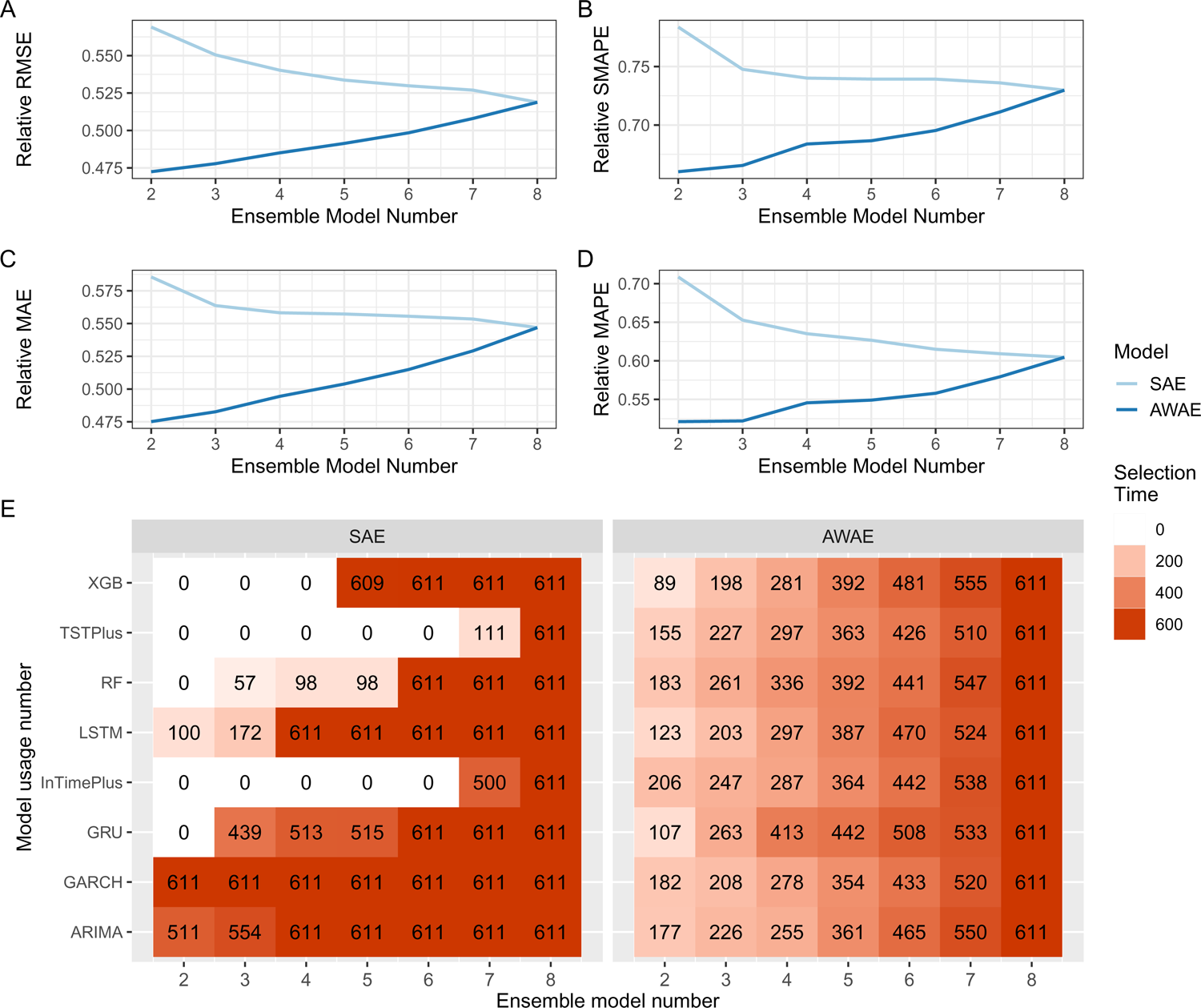

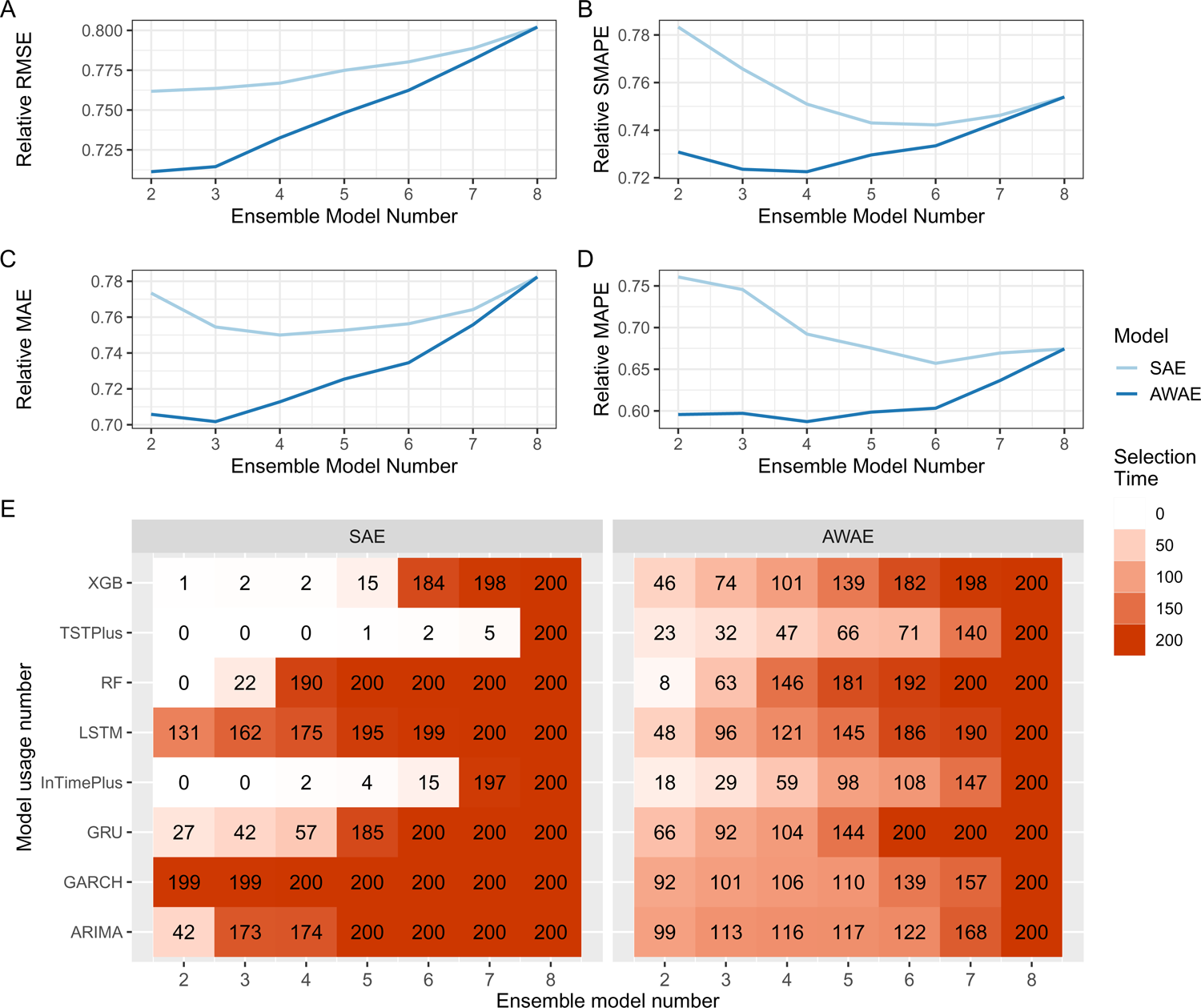

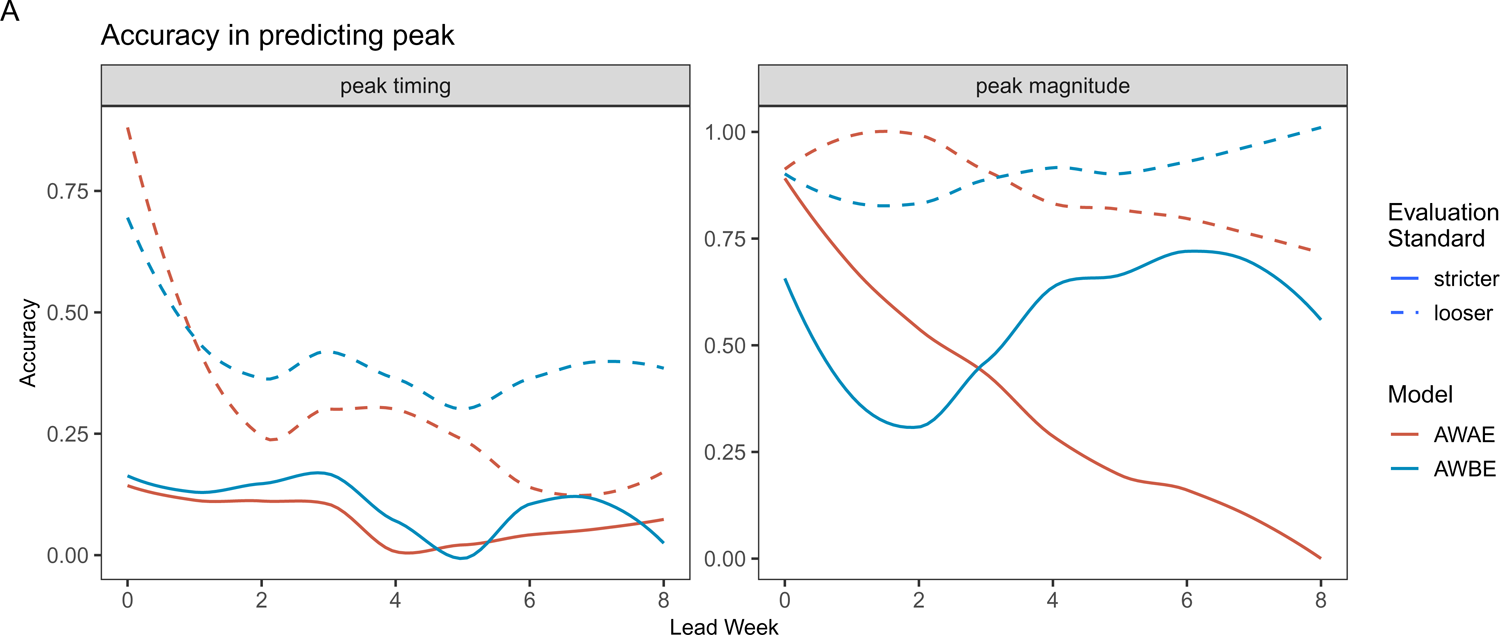

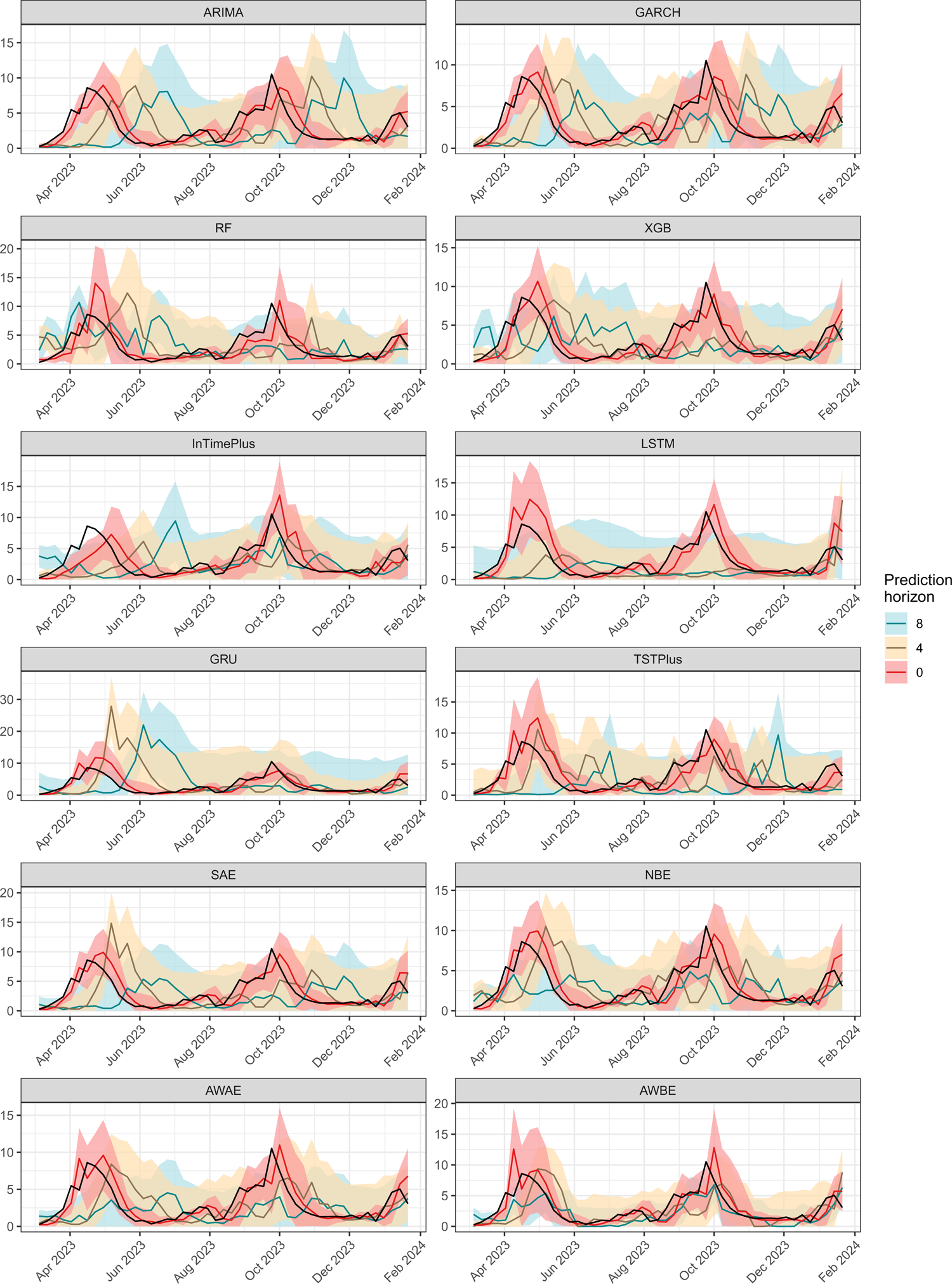

