## Supplementary Material for "An adaptive weight ensemble approach to forecast influenza activity in the context of irregular seasonality"

### Supplementary Material
### for ‘An adaptive weight ensemble approach to forecast influenza activity in the context of irregular seasonality’

### 1 Model details

#### 1.1 Description of individual models

##### 1.1.1 Baseline
The baseline model assumes that the ILI+ number stays at its current value indefinitely into the future, which means the most recently observed incidence $y_{t-1}$ is propagated forward to predict the incidence $y_{t+h}$.

##### 1.1.2 ARIMA
We fit a rolling ARIMA model to predict the ILI+ trend. The covariates are introduced in the model as lagged variables with lag $l_k$. The best lag for each covariate is estimated at each time step using the Pearson correlation coefficient between the ILI+ data and the covariate, with a maximum lag of 14. The parameters are estimated at each time step using the "auto.arima" function of the R package "forecast".

##### 1.1.3 GARCH
The Generalized Autoregressive Conditionally Heteroskedastic (GARCH) model is another traditional statistical model for time-series data. Here the model is fitted using the ugarchspec and ugarchfit function of the R package rugarch at each time step. The best lag for each covariate is estimated at each time step using the Pearson correlation coefficient between the ILI+ data and the covariate, with a maximum lag of 14. According to the grid search result, the hyperparameters order of arima part is set to be 14, and the order of garch part is set as 1.

### 1.1.4 RF
Random Forest (RF) is a bagging ensemble learning approach, which consists of several independent decision trees operating as a portfolio. The randomness that contributes to enhancing the accuracy and robustness of the model, comes from the use of bootstrap on features and training sets. Here the model is fitted based on the sklearn package in Python, and the best hyperparameters are determined through a grid search for each rolling. The covariates are introduced in the model as lagged variable series $[X_{t-1}, X_{t-2}, X_{t-k}]$ with lag k, after selected the optimal k as described in ARIMA.

##### 1.1.5 XGB
Extreme Gradient Boosting (XGB) model is a boosting ensemble learning approach. It obtains the final predictions by sequentially combining the results of multiple basic learning trees. In detail, a new tree that attempts to fit the residual error generated from the previous tree is built for each step. Contrary to RF, the individual trees included in XGB rely heavily on each other. The hyperparameters are confirmed through grid search for each rolling, using the Python package xgboost and sklearn. The covariates are also introduced in the model as lagged

variable series $[X_{t-1}, X_{t-2}, X_{t-k}]$ with lag k, after selected the optimal k as described in ARIMA.

##### 1.1.6 LSTM

The traditional sequential neural network, Long Short Term Memory network, which is a type of recurrent neural networks, can learn order dependence in sequence prediction problems [1]. It has the form of a chain of repeating modules of neural network, with the repeating module of four interacting neural network layer. Here we develop two ordinary LSTM stacked models, referred to as the Double_LSTM model, based on the actual acquisition sequence of ILI+ data. First, we build a one-horizon prediction LSTM, using the ILI+ and predictors up to week t-1, to make the nowcasting forecasting. Then we build the another eight-horizon prediction LSTM model, with the predictors up to week t and predicted week t ILI+, combined with historical ILI+ up to week t-1, to make forecasting for week t+1 – t+8. In this format, unlike the normal eight-horizon prediction LSTM, we can make maximum use of existing data, and the model shows better perfromance. Also, we compare the weighted MSE loss as well as the normal MSE loss for their forecasting performance to make the best model. As hyperparameter tuning is not the primary objective of this paper, the hyperparameters are confirmed through simple grid searching over training period and their values are as followings:

| process | hyperparameter | value | defnation |
|---|---|---|---|
| nowcasting | sequence_length | 14 | the length of sequence for model input |
| | batch_size | 6 | the size of one batch of data |
| | hidden_layers | 1 | the number of LSTM layers |
| | mid_dim | 72 | the middle dimension in the network, also the output dimension for single LSTM module |
| | dropout_rate | 0.3 | the dropout rate between LSTM layers |
| | num_directions | 1 | the direction of LSTM model, ot could be 1 or 2. |
| | num_epochs | 60 | the number of training epochs |
| | lr | 0.02 | the learning rate when training the model |
| forecasting | sequence_length | 14 | the length of sequence for model input |
| | batch_size | 6 | the size of one batch of data |
| | hidden_layers | 1 | the number of LSTM layers |
| | mid_dim | 72 | the middle dimension in the network, also the output dimension for single LSTM module |
| | dropout_rate | 0.3 | the dropout rate between LSTM layers |

| | | | |
|---|---|---|---|
| | num_directions | 1 | the direction of LSTM model, ot could be 1 or 2. |
| | num_epochs | 60 | the number of training epochs |
| | lr | 0.02 | the learning rate when training the model |

For LSTM data input, we set the size of a batch as 6 and the length of sequence as 14. For each LSTM model, the number of hidden layers is 1, the middle dimension is 72 and the direction is single direction. When training the model, the number of epochs is set to 60, with an original learning rate of 0.02. The learning rate will be adjusted by step based on the change of loss function.

##### 1.1.7 GRU

Gated Recurrent Neural Network (GRU), which is also a type of recurrent neural networks, is similar to LSTM model [2]. Compared with LSTM model, there are only three interacting neural network layers (two of them are update gate and reset gate separately) in each repeating module, thus making it run faster. The selection of hyperparameters and modeling format in the GRU is the same as LSTM model. The final values of hyperparameters are as follows:

| process | hyperparameter | value | defnation |
|---|---|---|---|
| nowcasting | sequence_length | 14 | the length of sequence for model input |
| | batch_size | 6 | the size of one batch of data |
| | hidden_layers | 1 | the number of LSTM layers |
| | mid_dim | 48 | the middle dimension in the network, also the output dimension for single LSTM module |
| | dropout_rate | 0.3 | the dropout rate between LSTM layers |
| | num_directions | 1 | the direction of LSTM model, ot could be 1 or 2. |
| | num_epochs | 50 | the number of training epochs |
| | lr | 0.04 | the learning rate when training the model |
| forecasting | sequence_length | 7 | the length of sequence for model input |
| | batch_size | 16 | the size of one batch of data |
| | hidden_layers | 1 | the number of LSTM layers |
| | mid_dim | 48 | the middle dimension in the network, also the output dimension for single LSTM module |
| | dropout_rate | 0.1 | the dropout rate between LSTM layers |
| | num_directions | 1 | the direction of LSTM model, ot could be 1 or 2. |

| | | | |
|---|---|---|---|
| | num_epochs | 50 | the number of training epochs |
| | lr | 0.0022 | the learning rate when training the model |

##### 1.1.8 TSTPlus

TSTPlus is a transformer-based time series framework model [3]. Unlike LSTM and GRU, TSTPlus model learns multivariate time series based on the transformer framework. In theory, Transformer uses a self-attention mechanism to process sequences, making it more efficient to process long sequences. The hyperparameters of TSTPlus, confirmed using grid searching, are as follows:

| process | hyperparameter | value | defnation |
|---|---|---|---|
| nowcasting | sequence_length | 14 | the length of sequence for model input |
| | d_model | 64 | the middle dimension in the network |
| | n_layers | 4 | the number of TSTEncode layer that to be combined |
| | dropout | 0.1 | the dropout rate between layers |
| | attention_dropout | 0.25 | The dropout rate for self-attention layers |
| | learning_rate | 0.0028 | the learning rate |
| | batch_size | 8 | the size of one batch of data |
| | n_epochs | 50 | the number of training epochs |
| forecasting | sequence_length | 14 | the length of sequence for model input |
| | d_model | 64 | the middle dimension in the network |
| | n_layers | 4 | the number of TSTEncode layer that to be combined |
| | dropout | 0.1 | the dropout rate between layers |
| | attention_dropout | 0.25 | The dropout rate for self-attention layers |
| | learning_rate | 0.0028 | the learning rate |
| | batch_size | 8 | the size of one batch of data |
| | n_epochs | 50 | the number of training epochs |

##### 1.1.9 InceptionTime Plus

InceptionTime Plus (InTimePlus) is an ensemble of deep Convolutional Neural Network (CNN) models, inspired by the Inception-v4 architecture [4]. The composition of an Inception network classifier contains two different residual blocks, as opposed to ResNet, which is comprised of three. For the Inception network, each block is comprised of three Inception modules rather than traditional fully convolutional layers. Each residual block's input is transferred via a shortcut linear connection to be added to the next block's input, thus mitigating the vanishing gradient problem by allowing a direct flow of the gradient [5]. InceptionTime model is an ensemble of 5 Inception networks, with each prediction given an even weight. The hyperparameters of InceptionTimePlus, confirmed using grid searching, are as follows:

| process | hyperparameter | value | defnation |
|---|---|---|---|
| nowcasting | sequence_length | 14 | the length of sequence for model input |
| | batch_size | 8 | the middle dimension in the network |
| | num_filters | 32 | the filter length of the Inception modules |
| | dropout_rate | 0.4 | the dropout rate between layers |
| | learning_rate | 0.0018 | the learning rate |
| | depth | 1 | the depth of layers |
| Forecasting | sequence_length | 14 | the length of sequence for model input |
| | batch_size | 6 | the middle dimension in the network |
| | num_filters | 32 | the filter length of the Inception modules |
| | dropout_rate | 0.2 | the dropout rate between layers |
| | learning_rate | 0.0024 | the learning rate |
| | depth | 5 | the depth of layers |

#### 1.2 Extra supplements of individual models

##### 1.2.1 Rolling method

We roll the forecast origin to generate the forecasting results. For each week t, we forecast ILI+ for week t ~ t+8 using meterological data up to week t (since the mererolgoical data is available in daily basis), and the ILI+ data which is only up to week t-1. In real-time, for week t, the ILI+ is only available up to week t-1 but the predictors are available up to week t, so we design this form of prediction.

For the machine learning models, both weekly rolling and yearly rolling methods are tested. We conducted experiments for GRU and LSTM in testing period. Compared to the annual rolling window method, the weekly rolling window approach requires much more computational times; however, the discrepancies in accuracy for individual models were minimal. Intriguingly, the individual models trained utilizing the weekly rolling window approach may slightly worse the prediction performance of the ensemble model (Figure S1). Therefore, when finally building the simulated ensemble forecast framework, we adopt the less time-consuming year-rolling modeling approach for machine learning models.

#### 1.3 Description of ensemble models

The differences in the performance of various individual models motivate us to try ensemble models.

##### 1.3.1 Simple Average Ensemble

Firstly we test the Simple Average Ensemble (SAE) model. This model is created by directly selecting the three best-performing models for unweighted average forecasting based on their RMSE scores. Model ensembling is also conducted in a rolling manner, which means that we select the top three models every week and use their average value as the ensemble result.

##### 1.3.2 Normal Blending Ensemble

We also try the blending ensemble approach, in which we use LASSO regression without the intercept term to automatically select models and determine their weights. For each week t, the LASSO regression is fitted over the full historical data, and then the coefficients are applied to generate the weighted ensemble predictions. The regularization coefficient parameter α is determined using a five-fold cross-validation at each week.

##### 1.3.3 Adaptive Weighted Ensemble

The Adaptive Weighted Ensemble models are inspired by the time decay and sample weighting ideas. For each week t, we first weigh the different samples with the exponential decay function: $w_{t-k} = e^{-\lambda \cdot k}$, and then use the weighted samples to make ensemble. For the Simple Averaging Ensemble (SAE) model, we first apply sample weights to the RMSE and then select the top three models based on the weighted RMSE indicators. The weighted RMSE for each model is calculated as follows:

$$RMSE_{m,t} = \sqrt{\frac{1}{t}\sum_{i=1}^{t-1} w_i (y_i - \widehat{y_i^m})^2}, \qquad w_i = e^{-\lambda \cdot (t-i)},$$

where $y_i$ means the true ILI+ vector at time i, $\widehat{y_i^m}$ means the predicted ILI+ vector, $m$ represents the model and $\lambda$ represents the time decay hyperparameter which equals 0.1 here.

For the blending ensemble method, we fit a weighted LASSO model during the LASSO regression by using time decay coefficients as sample weights, and then perform ensemble prediction based on the obtained coefficients. The loss function for this weighted LASSO is:

$$L_t = \sum_{i=1}^{t-1}\left( w_i \cdot \left( y_i - \sum_{m=1}^{M} \beta_m \widehat{y_i^m} \right)^2 + \delta \sum_{m=1}^{M} |\beta_m| \right), w_i = e^{-\lambda \cdot (t-i)},$$

where $m$ represents the model and t represents the time t. We set the time decay rate $\lambda$ is 0.1.

### 2 Predictors

We include a series of predictors in individual models. Two types of predictors are considered in our study, 12 epidemiological predictors and 8 meteorological predictors. The best lag of these covariates is estimated at each time step using the Pearson correlation coefficient between the covariates and the dependent variable. The maximum lag is fixed and set to be 14 for all predictors.

#### 2.1 Epidemiological predictors

Besides current $Y_t$ , we consider that the past temporal Y may be correlated with $Y_t$, especially the past two weeks. Hence, we explored temporal $R_{t-m}^{t}$ (m = 0, ..., 10)in the past 10 days. As the influenza seasonality in HK is well documented (a winter peak and summer peak) [6], so the week number in a year and the month in a year are included in all models.

#### 2.2 Meteorological predictors

Climate data are obtained from Hong Kong Observatory (hko.gov.hk), including pressure, temperature, relative humidity, amount of cloud, rainfall, number of hours of reduced visibility, total bright sunshine, global solar radiation, evaporation, and wind speed. Due to high correlations among these variables, we finally select meteorological predictors, including the maximum and the minimum of temperature, the relative humidity, total rainfall, and solar radiation based on correlation and previous literatue[7].

### 3 Model Evaluation

#### 3.1 Evaluation Matrix

A total of four evaluation indicators are referenced, including RMSE [8], SMAPE [9, 10], WIS [11], MAE [12] and MAPE. Let $e_i = \widehat{y_i} - y_i$ represent the bias error for each sample $i$, then the calculation formula is as follows:

$$RMSE = \left[\frac{1}{n} \cdot \sum_{i=1}^{n} |e_i|^2\right]^{\frac{1}{2}},$$

$$SMAPE = \frac{1}{n}\sum_{i=1}^{n} \frac{|e_i|}{(|\widehat{y_i}| + |y_i|)},$$

$$WIS_{\alpha_{0:K}}(F,y) = \frac{1}{K+1/2} \cdot \left( w_0 \cdot |y-m| + \sum_{k=1}^{K} w_k \cdot IS_{\alpha_k}(F,y) \right), where\ IS_{\alpha}(F,y)$$

$$= (u-l) + \frac{2}{\alpha} \cdot (l-y) \cdot 1\{y < l\} + \frac{2}{\alpha} \cdot (y-u) \cdot 1\{y > u\},$$

$$MAE = \frac{1}{n}\sum_{i=1}^{n} |e_i|,$$

$$MAPE = \frac{1}{n}\sum_{i=1}^{n} \frac{|e_i|}{|y_i|}.$$

To facilitate a more comprehensive assessment and comparison of the models, we compute the relative metrics for each model, which indicate its performance in relation to the baseline model. Using RMSE as an illustration, the corresponding equation is presented below:

$$RMSE^{m}_{relative} = \frac{RMSE^{m}_{true}}{RMSE^{baseline}_{true}}, \qquad for\ Model\ m.$$

#### 3.2 Interval Forecast

Deep learning models typically focus on point predictions, and a prevalent approach for generating interval forecasts which is called Monte Carlo Dropout (MC Dropout) involves using dropout layers in neural networks to account for uncertainty in deep learning, thereby producing interval predictions [13, 14]. Our experiment suggests that although MC Dropout enables the generation of interval forecasts, it may worse the point prediction performances (Figure S2). Consequently, we do not use MC Dropout in our study.

To tackle the aforementioned issue, we employed a Gaussian distribution-based interval forecasting approach [15]. This method assumes that the prediction outcomes follow a normal distribution, with the mean corresponding to the predicted value $\hat{y}$ and the standard deviation derived from the prediction residuals within a specific rolling window $k$. We relaxed the constraint of the window length $k = 20$ and calculated the corresponding standard deviations $\sigma_k$ and interval prediction results for various window lengths $k$ within the 5-50 range. We determined the optimal window length $k_h$ for each prediction horizon $h$, by selecting the value k so that the proportion of coverage of the true value of 90% prediction interval is close to 90%, based on the training set. Subsequently, we generated the final interval prediction outcomes on the test set using these optimal window lengths.

### 4 Supplementary Tables

#### 4.1 Supplementary Table 1 Model performance in comparison to Baseline

**Table 1. Model performance in comparison to naive persistence baseline.** Each cell represents the relative value of evaluation metrics for individual and ensemble models compared to baseline model metrics, inclucing RMSE, SMAPE, WIS, MAE and MAPE. Values less than 1 indicate improvement over the baseline model, with smaller values representing greater improvement.

| | | RMSE | | | | SMAPE | | | | WIS | | | | MAE | | | | MAPE | | | |
|---|---|---|---|---|---|---|---|---|---|---|---|---|---|---|---|---|---|---|---|---|---|
| | | Week0 | Week2 | Week4 | Week8 | Week0 | Week2 | Week4 | Week8 | Week0 | Week2 | Week4 | Week8 | Week0 | Week2 | Week4 | Week8 | Week0 | Week2 | Week4 | Week8 |
| Individual Models | ARIMA | 1.00 | 0.83 | 0.76 | 0.67 | 0.97 | 0.83 | **0.78** | 0.73 | 0.98 | 0.80 | 0.74 | 0.67 | 0.97 | 0.77 | 0.70 | 0.62 | 1.01 | 0.87 | 0.77 | 0.58 |
| | GARCH | **0.94** | 0.82 | 0.76 | 0.67 | **0.94** | **0.85** | 0.82 | 0.79 | **0.91** | 0.81 | 0.75 | 0.69 | 0.93 | 0.79 | 0.73 | 0.67 | 0.99 | 0.96 | 0.91 | 0.80 |
| | RF | 1.21 | **0.79** | **0.69** | **0.63** | 1.12 | 0.87 | 0.79 | 0.74 | 1.17 | **0.71** | **0.66** | **0.64** | 1.11 | 0.78 | 0.67 | 0.63 | 1.09 | 0.83 | 0.68 | 0.53 |
| | XGB | 1.09 | 0.86 | 0.72 | 0.66 | 1.06 | 0.92 | 0.82 | 0.76 | 1.06 | 0.78 | 0.70 | 0.67 | 1.04 | 0.85 | 0.72 | 0.66 | 1.07 | 0.93 | 0.75 | 0.57 |
| | InTimePlus | 1.43 | 0.84 | 0.78 | **0.63** | 1.33 | 0.91 | 0.80 | **0.69** | 1.39 | 0.81 | 0.75 | **0.62** | 1.36 | 0.85 | 0.75 | 0.60 | 1.50 | 0.98 | 0.82 | 0.47 |
| | TSTPlus | 1.16 | 0.85 | 0.73 | 0.65 | 1.10 | 0.93 | 0.82 | **0.70** | 1.14 | 0.81 | 0.71 | 0.65 | 1.10 | 0.85 | 0.72 | 0.63 | 1.23 | 1.13 | 0.80 | 0.51 |
| | LSTM | **0.92** | 0.81 | 0.75 | 0.67 | **0.91** | 0.86 | **0.78** | 0.75 | **0.87** | 0.80 | 0.74 | 0.67 | 0.88 | 0.81 | 0.73 | 0.66 | 0.95 | 0.90 | 0.71 | 0.55 |
| | GRU | 1.06 | **0.79** | **0.70** | 0.65 | 0.99 | **0.84** | **0.77** | 0.74 | 0.99 | **0.72** | **0.69** | 0.65 | 0.98 | 0.78 | 0.69 | 0.65 | 0.96 | 0.83 | 0.70 | 0.55 |
| Ensemble Models | SAE | 0.94 | 0.77 | 0.70 | 0.64 | 0.91 | 0.78 | 0.75 | 0.72 | 0.90 | 0.76 | 0.70 | 0.65 | 0.90 | 0.73 | 0.66 | 0.61 | 0.93 | 0.83 | 0.75 | 0.62 |
| | NBE | 0.93 | 0.81 | 0.71 | 0.63 | 0.94 | 0.86 | 0.80 | 0.78 | 0.90 | 0.75 | 0.72 | 0.67 | 0.93 | 0.81 | 0.72 | 0.67 | 1.02 | 1.09 | 0.97 | 0.84 |
| | AWAE | **0.88** | **0.71** | 0.62 | 0.54 | **0.89** | **0.76** | **0.68** | 0.60 | **0.87** | **0.70** | 0.60 | 0.53 | 0.88 | 0.69 | 0.58 | 0.49 | 0.91 | 0.77 | 0.62 | 0.44 |
| | AWBE | 1.05 | 0.79 | **0.50** | **0.42** | 0.97 | 0.89 | **0.70** | **0.54** | 0.97 | 0.75 | **0.50** | **0.42** | 0.98 | 0.74 | 0.50 | 0.39 | 0.95 | 0.96 | 0.60 | 0.35 |

#### 4.2 Supplementary Table 2 Model Performance for the Post-COVID-19 Pandemic Rebound

**Table 2. Average Model Performance Metrics (0-8 Prediction Horizon) Compared to Naive Persistence Baseline for 2023.** Each cell in the table displays the ratio of the model's metric values to the corresponding persistence model metric values; a value less than 1 signifies an improvement in the model compared to the persistence method.

| | | RMSE | SMAPE | WIS | MAPE | MAE |
|---|---|---|---|---|---|---|
| Individual model | ARIMA | 1.00 | 0.99 | 0.99 | **0.86** | 0.98 |
| | GARCH | 0.88 | 0.91 | 0.87 | **0.78** | 0.86 |
| | RF | 0.95 | 0.92 | 0.96 | 1.56 | 0.92 |
| | XGB | **0.85** | **0.89** | **0.84** | 1.04 | **0.82** |
| | InTimePlus | **0.83** | **0.90** | **0.83** | 0.88 | 0.79 |
| | TSTPlus | 0.91 | 1.04 | 0.88 | 0.91 | 0.89 |
| | LSTM | 0.91 | 1.06 | 0.85 | 0.83 | **0.84** |
| | GRU | 1.36 | 0.93 | 1.20 | 1.27 | 1.12 |
| Ensemble model | SAE | 0.92 | 0.94 | 0.90 | 0.76 | 0.89 |
| | NBE | 0.79 | **0.86** | **0.77** | **0.69** | 0.77 |
| | AWAE | **0.76** | **0.85** | 0.78 | 1.00 | **0.76** |
| | AWBE | **0.68** | 0.88 | **0.64** | **0.57** | **0.62** |

#### 5 Supplementary Figure Legend

**Figure S1.** A comparison of prediction performance and time consumption between yearly rolling and weekly rolling modeling approaches. Panel A: Performance of yearly and weekly rolling models, including LSTM, GRU, AWAE, and AWBE. Panel B: Time consumption comparison for LSTM and GRU models.

**Figure S2.** Comparison of model performance based on different MC Dropout rates. Panel A: RMSE; Panel B: SMAPE; Panel C: MAPE; Panel D: MAE.

**Figure S3.** The trajectory of the models' forecasting outcomes. The featured models include all the individual models and ensemble models.

**Figure S4.** Interval trajectory of the models' forecasting outcomes, featuring all the individual and ensemble models. Different colors represent various prediction horizons.

**Figure S5**. A comparative analysis of Simple Average Ensemble and Adaptive Weighted Average Ensemble performance with varying numbers of individual models (1-8) in model ensemble. Panel A: RMSE. Panel B: SMAPE. Panel C: MAE. Panel D: MAPE. Panel E: In the ensemble model, when selecting different numbers of individual models, the frequency of each individual model being chosen was recorded. A maximum value of 200 implies that the entire training dataset comprises 200 weeks of data, indicating that a total of 200 model ensembles were performed on the training set.

**Figure S6.** Relative RMSE and time consumption of various models across differing training data lengths. The relative RMSE is calculated in comparison to the Baseline model. The ensemble models' time consumption comprises the sum of all individual model durations and the time cost of ensemble implementation.

**Figure S7.** Accuracy in predicting outbreak peak timing and peak magnitude. Results are shown for AWAE (red) and AWBE (blue), evaluated using two standards (solid vs. dashed lines, as specified in the legend).

**Figure S8.** Interval trajectory of the models' forecasting outcomes during the post-COVID period, featuring both individual and ensemble models. Distinct colors represent various prediction horizons.
